# A maternal germline mutator phenotype in a family affected by heritable colorectal cancer

**DOI:** 10.1101/2023.12.08.23299304

**Authors:** Candice L. Young, Annabel C. Beichman, David Mas-Ponte, Shelby L. Hemker, Luke Zhu, Jacob O. Kitzman, Brian H. Shirts, Kelley Harris

**Affiliations:** Department of Genome Sciences, University of Washington, 3720 15th Ave NE, Seattle, WA 98195; Department of Molecular and Cellular Biology, University of Washington, 1705 NE Pacific St, Seattle, WA 98195; Department of Human Genetics, University of Michigan, 1241 Catherine St, Ann Arbor, MI 48109; Department of Bioengineering, University of Washington, 3720 15th Ave NE, Seattle, WA 98195; Department of Laboratory Medicine and Pathology, University of Washington, 1959 NE Pacific St, Seattle, WA 98195; Herbold Computational Biology Program, Fred Hutchinson Cancer Center, P.O. Box 19024, Seattle, WA 98109

**Keywords:** germline mutagenesis, de novo mutation calling, mutational signature, MUTYH-associated polyposis, mutator allele, oxidative damage, pedigree

## Abstract

Variation in DNA repair genes can increase cancer risk by elevating the rate of oncogenic mutation. Defects in one such gene, *MUTYH*, are known to elevate the incidence of colorectal cancer in a recessive Mendelian manner. Recent evidence has also linked *MUTYH* to a mutator phenotype affecting normal somatic cells as well as the female germline. Here, we use whole genome sequencing to measure germline de novo mutation rates in a large extended family containing both mothers and fathers who are affected by pathogenic *MUTYH* variation. By developing novel methodology that uses siblings as “surrogate parents” to identify de novo mutations, we were able to include mutation data from several children whose parents were unavailable for sequencing. In the children of mothers affected by the pathogenic *MUTYH* genotype p.Y179C/V234M, we identify an elevation of the C>A mutation rate that is weaker than mutator effects previously reported to be caused by other pathogenic *MUTYH* genotypes, suggesting that mutation rates in normal tissues may be useful for classifying cancer-associated variation along a continuum of severity. Surprisingly, we detect no significant elevation of the C>A mutation rate in children born to a father with the same *MUTYH* genotype, and we similarly find that the mutator effect of the mouse homolog *Mutyh* appears to be localized to embryonic development, not the spermatocytes. Our results suggest that maternal *MUTYH* variants can cause germline mutations by attenuating the repair of oxidative DNA damage in the early embryo.

## Introduction

Many DNA repair deficiencies are linked with increased risk for cancer syndromes (Fearon 1997; Goode et al. 2002; Matullo et al. 2006; Randall et al. 2023). Pathogenic mutations leading to the loss of function in specific DNA repair pathways accelerate the accumulation of oncogenic variants. While each DNA repair defect often tends to cause cancers mainly in specific tissues, other tissues may also accumulate a higher mutation load than normal (Scarbrough et al. 2016; Dunlop et al. 1997; Aarnio et al. 1999). It is not well understood why accelerated mutagenesis only seems to lead to cancer in certain tissues, or whether somatic mutations that do not cause cancer might have other health impacts (Blokzijl et al. 2016; Elledge and Amon 2002; Chao and Lipkin 2006).

Some recent studies (Sherwood et al. 2023; Andrianova et al. 2023; Stendahl et al. 2023; Kaplanis et al. 2022) have paid particular attention to the impact of DNA repair deficiencies on the germline because even modestly elevated germline mutation rates can impact congenital disease risk and the rate of evolution. Moreover, since germline mutations can be studied through relatively straightforward comparisons between relatives (Wei et al. 2015; Bergeron et al. 2022) and do not require the specialized technologies that are needed to detect low-frequency somatic variants (Kennedy et al. 2014; Ellis et al. 2021), germline mutator phenotypes have the potential to lead to discovery of new DNA repair defects that may be candidate drivers of novel cancer syndromes. For example, inherited variation was recently used to discover that a variant in the murine Base Excision Repair (BER) DNA-glycosylase *Mutyh* gene acts as a germline mutator allele in inbred mouse strains (Sasani et al. 2022, 2023). Since impaired functioning of the human MUTYH protein is known to cause a colorectal cancer syndrome known as *MUTYH*-associated polyposis (MAP) (Smith et al. 2013a), this mutator allele is a promising candidate for exploring joint effects of DNA repair genes on the mammalian soma and germline.

The *MUTYH* gene plays a key role in BER, a DNA repair pathway that evolved to repair damage caused by reactive oxygen species (ROS), which are byproducts of aerobic metabolism (Banda et al. 2017). ROS can react with guanine to create the lesion 8-oxoguanine (8-OG), which has a propensity to mispair with adenine, resulting in G:C > T:A transversion mutations, often abbreviated as C > A mutations (David et al. 2007). BER DNA glycosylases have developed a specific mechanism to repair this mutagenic damage: OGG1 removes 8-OG from the compromised strand (Hayashi et al. 2002) while MUTYH excises the erroneously incorporated adenines opposite 8-OG (Woods et al. 2016; Krokan and Bjørås 2013). Due to MUTYH*’*s role in this repair pathway, defects in this enzyme can cause excess accumulation of C>A mutations in tissues that are experiencing ROS damage (Pilati et al. 2017).

*MUTYH*-associated polyposis (MAP) follows a recessive inheritance pattern, affecting individuals who have inherited two sub-functional copies of the *MUTYH* gene (Morak et al. 2014), causing intestinal adenomatous polyposis as well as an elevated risk for early-onset colorectal and duodenal malignancies (Nielsen et al. 2011; Al-Tassan et al. 2002). The genotypes that cause MAP are often referred to as “biallelic”: biallelic *MUTYH* genotypes are either homozygous for a single pathogenic variant or are compound heterozygotes, with each copy of *MUTYH* affected by a different pathogenic mutation. In contrast, individuals who have “monoallelic” genotypes (meaning heterozygous for a single pathogenic *MUTYH* variant) do not generally develop intestinal polyposis and have at most a modestly elevated cancer risk (Barreiro et al. 2022). Notably, *MUTYH* is an example of a gene that plays a crucial role in genomic stability across all tissues affected by ROS damage, but mainly appears to modulate cancer risk in the colorectal epithelium (Nieuwenhuis et al. 2012; Hutchcraft et al. 2021). Despite the tissue specificity of MAP’s cancer risk phenotype, recent evidence indicates that this condition also causes elevated somatic mutation rates in a wider variety of human cell types, including blood (Robinson et al. 2022), which might be why some studies have found *MUTYH* variants to be associated with increased risk of extracolonic cancers (Vogt et al. 2009; Win et al. 2016; Zhang et al. 2006; Beiner et al. 2009; Villy et al. 2022). These findings led us to hypothesize that *MUTYH*’s C>A mutator effect might extend to germline cells.

To test whether pathogenic *MUTYH* genotypes might cause a human germline mutator phenotype, we sequenced fifteen genomes from a large extended family containing multiple individuals affected by MAP. We used these genomes to test whether parent/child trios including a parent with MAP show evidence of elevated C>A mutation rates compared to trios where the parental *MUTYH* genotypes are normal or monoallelic. Recently, another group published de novo mutation data from two trios whose mothers were affected by MAP (Sherwood et al. 2023), obtaining evidence for a C>A mutator effect. We further investigated this effect in our larger dataset, which includes children of both mothers and fathers affected by the same pathogenic *MUTYH* genotype. We then further contextualized our results through a comparison to a null model of mutation rate as function of parental age that was previously constructed from thousands of control trios (Jónsson et al. 2017). In this way, we were able to characterize how pathogenic *MUTYH* variants affect the human germline, using an analysis framework that is broadly appropriate for investigating the effects of other cancer syndromes on germline mutagenesis and human evolution.

## Results

### Whole genome sequencing of families affected by a MUTYH genotype that shows reduced DNA repair efficiency in vitro

We obtained saliva samples from three siblings (labeled P1, P2, and P3 in **Figure 1**) who share a biallelic *MUTYH* genotype known as p.Y179C/V234M. (One gene copy has a tyrosine-to-cysteine substitution at amino acid position 179 and the other has a valine-to-methionine substitution at position 234. Both amino acid positions are indexed in the coordinates of *MUTYH* transcript NM_001128425; substitutions p.Y179C and p.V234M have DNA coordinates c.536A>G and c.700G>A, respectively.) We also sampled saliva from a fourth sibling (P4 in **Figure 1**) who is a monoallelic carrier of p.V234M. These siblings inherited p.Y179C from their father and inherited p.V234M from their mother. Two of the three biallelic siblings were previously diagnosed with colon cancer, and the third has a history of colon polyps (all three meet the diagnostic criterion for MAP by virtue of their *MUTYH* genotype). The siblings’ extended family is broadly affected by an elevated colorectal cancer incidence. All sampled individuals tested negative for Lynch syndrome variants and other variants known to cause heritable colorectal cancer.

**Figure 1.**
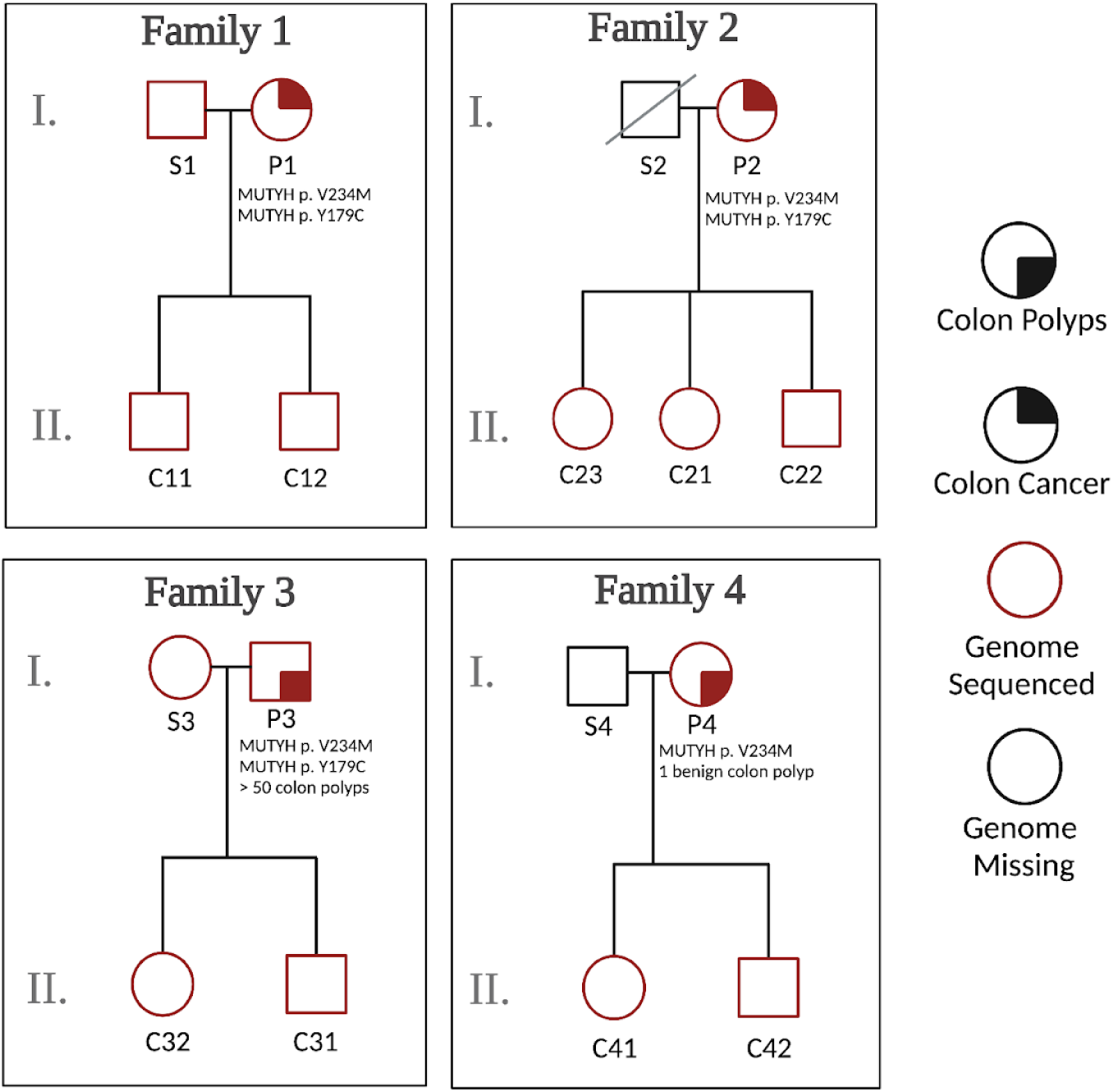
Sequencing four families of *MUTYH* variant carriers. To measure the effects of biallelic *MUTYH* mutations on germline mutagenesis, we sequenced three individuals with the same pathogenic *MUTYH* genotype, as well as one related monoallelic *MUTYH* variant carrier, along with their children and partners. Individuals have been given labels that indicate which nuclear family they are part of (1–4), whether they are a *MUTYH* variant carrier parent (P), a spouse or partner of that parent (S), or a child (C). Families 1 and 2 include mothers with the biallelic genotype p.Y179C/V234M, while Family 3 includes a father with the same p.Y179C/V234M genotype. Family 4 includes a mother with the monoallelic mutation p.V234M. Shaded quadrants indicate which individuals have been diagnosed with colon polyps (bottom right) or colon cancer (top right). *MUTYH* mutations and age at cancer diagnosis / number of identified colon polyps are listed below individuals for which this information is known. Square = male; circle = female; red = genome sequenced; black = genome not sequenced.

While ClinVar classifies p.Y179C as pathogenic with evidence from many previous studies (Al-Tassan et al. 2002; Nielsen et al. 2005, 2009; Vogt et al. 2009), some laboratories consider p.V234M to be a variant of uncertain significance with mixed functional evidence (Peterlongo et al. 2006; Fleischmann et al. 2004; Yurgelun et al. 2015; Komine et al. 2015). To obtain more information about the pathogenicity of this genotype p.Y179C/V234M, we conducted functional assays in which mutant *MUTYH* expression is restored in human HEK293 *MUTYH* KO cells. Our approach uses a reporter construct engineered to contain an 8-oxoG:A lesion, such that proper repair corrects a premature stop codon in GFP and restores its expression. Notably, the p.Y179C allele exhibited severe loss of repair function, whereas the p.V234M variant displayed a partial loss of function with repair activity well below that of wild-type MUTYH (**Figure S1**). The deleterious effects observed for these two variants within the HEK293 cell context indicate that they likely have pathogenic effects and may result in elevated mutation accumulation across tissues *in vivo*.

We also used the same functional assay to measure the effects of c.461GT>AA p.R182Q (corresponding to the DNA substitution c.461 GT>AA on transcript NM_001128425), the human analog of the mutation found in an outlier mouse strain known as BXD68 that displayed a *Mutyh* hypermutator germline phenotype (Sasani et al. 2022). p.R182Q appears to be a total loss of function variant with a phenotype similar to that of p.Y179C.

We note that the previous study of *MUTYH*’s germline mutator activity by Sherwood et al. (2023) analyzed families with a different compound heterozygous genotype known as p.Y179C/G368D. p.G368D (corresponding to the DNA substitution c. 1187 G>A on transcript NM_001128425) may be less deleterious than p.Y179C given its association with an older age at MAP diagnosis (Guarinos et al. 2014) and its less severe somatic mutator phenotype (Robinson et al. 2022). Note that Robinson, et al. refer to the variant p.G368D as p.G396D in the coordinates of a different reference *MUTYH* transcript.

To assess the impact of the *MUTYH* p.Y179C/V234M genotype on the germline mutation rate and spectrum, we performed 50X coverage whole genome sequencing on P1–P4 as well as nine of their adult children and two of their spouses (**Figure 1**). Individuals have been given labels according to which nuclear family they are a member of (1–4), and whether they are a *MUTYH* variant carrier parent (P), a spouse or partner of that parent (S), or a child of a *MUTYH* variant carrier (C). Colloquially, we refer to all parents as mothers and fathers if they conceived their children via oocytes and spermatocytes, respectively, although we recognize that these labels may not match parents’ individual gender identities.

### Using siblings as “surrogate parents” for de novo mutation calling in incomplete nuclear families

DNMs are typically called by identifying sites that violate the principles of Mendelian inheritance. These are sites at which a child’s genome contains a variant not observed in the genome of either of their parents. DNM calling normally requires the genomes of both parents to be sequenced, and Families 1 and 3 are the only nuclear families in our dataset that meet this requirement (**Figure 1**). Families 2 and 4 were not suitable for standard mutation calling due to unavailability of the paternal genomes (one father is deceased and the other declined to participate). To maximize the power of this study, we developed a novel method that facilitated DNM calling in these families. Although our new method has slightly lower accuracy and precision than standard DNM calling, particularly in Family 4 which consists of just a mother and two children, it enables DNM analysis in previously inaccessible families within our study and potentially beyond.

Since each child in generation II has at least one full sibling represented in our dataset, we were able to leverage the sharing of parental haplotypes among siblings to devise a “surrogate parent” method for estimating DNM rates and spectra. Instead of comparing each child’s genome to their mother and father to identify variants that must have arisen de novo, we compared each child from Families 2 and 4 to their mother plus one or more siblings who inherited some of the same paternal DNA. If the child’s genome contains a variant that is absent from both the mother and the sibling surrogate father, this implies that the unique variant arose de novo in the child (**Figure 2A-2B**; pipeline described in **Figure S2**). Jónsson, et al. (2018) previously used a similar procedure to identify mutations that arose early in parental embryonic development, which often have parental read support and are thus not detectable by standard parent/child trio mutation calling methodology.

**Figure 2.**
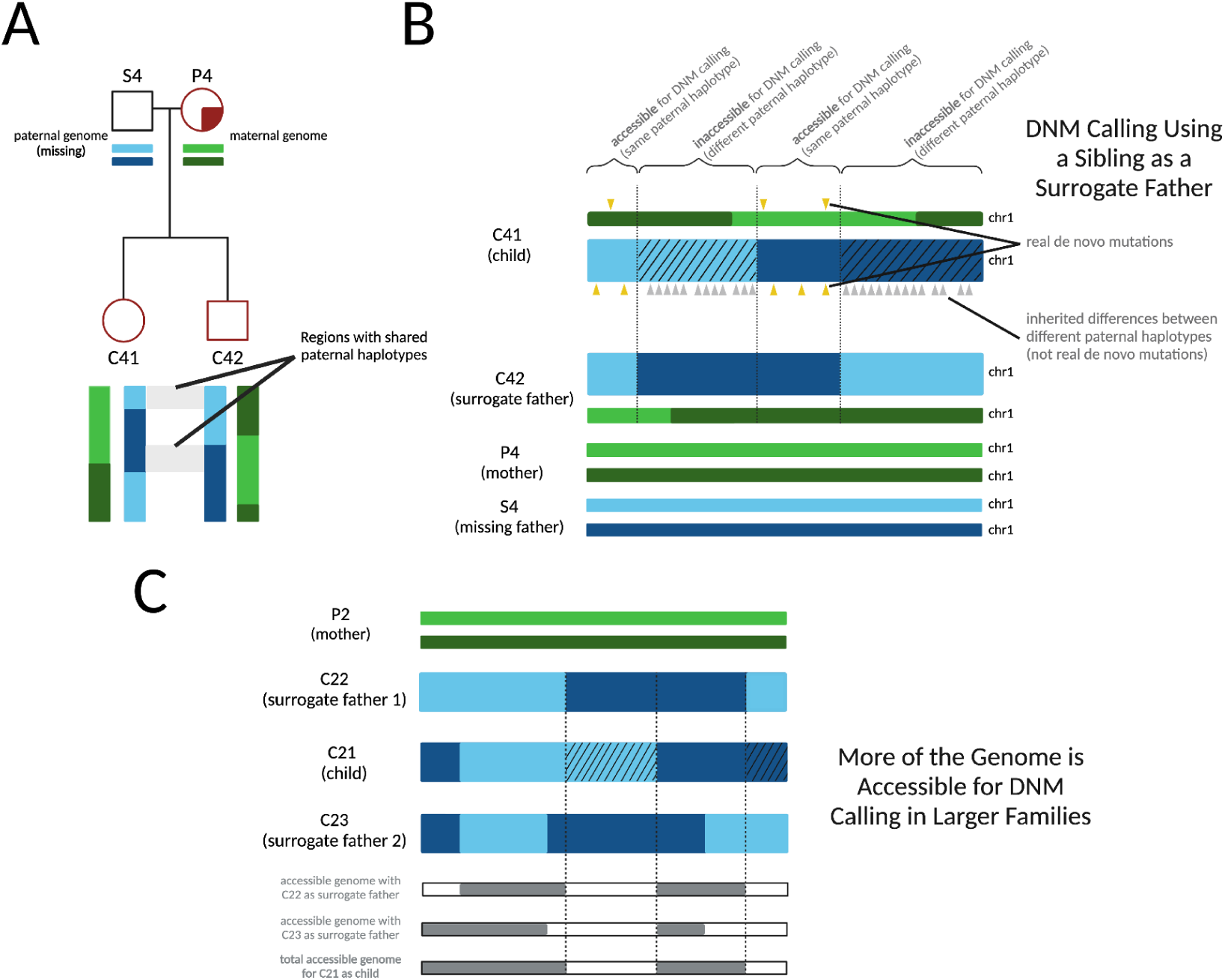
Calling DNMs using one parent and one surrogate parent. **A)** An illustration of the portions of an autosome with paternal haplotypes shared between two siblings. In an example chromosome from Family 4, DNMs can be called in regions where C41 and C42 share a paternal haplotype sequence with one another. **B)** An illustration of DNM calling using a sibling as surrogate father. In regions where the siblings inherited the same paternal haplotype, Mendelian violations (DNM calls, yellow triangles) are spaced far apart, but in regions where the siblings inherited different paternal haplotypes, Mendelian violations (gray triangles) are clustered close together, mostly stemming from polymorphic differences between the different paternal haplotypes inherited by the respective siblings. Hashed chromosome regions represent inaccessible regions of the genome, where DNMs cannot be called using the surrogate approach. **C)** An example of the surrogate method applied to Family 2, a three-child family where two different surrogate fathers can be used to call DNMs in each child. A set of partially overlapping candidate DNMs is generated from each sibling comparison, increasing the amount of accessible genome where mutations can be identified with more siblings used in this approach and allowing additional validation of calls in regions where accessible regions overlap.

As in standard DNM calling pipelines (Bergeron et al. 2022), we only call mutations at sites where both the real and surrogate parents are homozygous for the reference allele. This allows us to polarize mutation calls with confidence: if we are calling DNMs in Sibling 1 using Sibling 2 as a surrogate parent, all DNM calls will occur at sites where Sibling 1 is heterozygous and Sibling 2 is homozygous, which are not variants that could also be explained by DNMs in Sibling 2.

Our use of the term “surrogate parent” is loosely based on the established use of relatives as surrogate parents for haplotype phasing (Kong et al. 2008). We also drew inspiration from several recent studies that successfully estimated mutation rates using tracts inherited identical-by-descent (IBD) between relatively distant relatives (Narasimhan et al. 2017; Tian et al. 2019, 2022). In contrast to these earlier studies, which estimated population-averaged mutation rates using mutations that likely occurred many generations ago, our method is designed to estimate an individual parent/child trio’s mutation rate using mutations that arose within a single generation. This method enabled us to estimate germline mutation rates within five nuclear subfamilies of the pedigree and study how *MUTYH* genotype affected the germline mutation rate and spectrum.

To identify regions accessible for DNM calling using a mother and a surrogate father, we first used the software hap-IBD (Zhou et al. 2020) to identify long haplotypes shared identical by descent between each pair of siblings. We then filtered these regions to exclude maternally inherited haplotypes. In addition, we implemented a “SNP density filter” to exclude what appear to be false positive IBD calls: regions where the density of pairwise differences between the siblings is too high to be consistent with true sharing of paternal haplotypes (see Methods). Within the remaining regions of paternal haplotype sharing, we called DNMs using a standard GATK-based pipeline, using the surrogate father as the father. In this setup, gene conversion between the two paternal haplotypes has the potential to create false positive DNMs, as previously observed by Narasimhan et al. (2017). To minimize these errors, we filtered out putative DNMs that were present in the 1000 Genomes data or in two or more members of our pedigree (this should eliminate gene conversion errors at any loci where not all siblings inherited the same paternal haplotype). We note that the familial mutation sharing filter will cause us to miss the 1-2% of DNMs that are shared between siblings due to germline mosaicism (Jónsson et al. 2018). The 1000 Genomes filter may also cause us to exclude some true DNMs, particularly at CpG sites or other mutation hotspots, but it should effectively filter out many false positive DNMs that were actually inherited from a missing parent, except when those inherited variants are very rare in the population as a whole.

In a large family with a mother and *n*+1 siblings, the only regions inaccessible for surrogate-father DNM calling will be regions where *n* siblings inherited the same paternal haplotype and the remaining sibling inherited the other paternal haplotype. In this scenario, the sibling who inherited the unique paternal haplotype has no one to serve as a surrogate father, so that sibling’s genome will be inaccessible for DNM calling while the *n* other siblings’ genomes will be accessible. If we consider the paternal haplotype inherited at a specific locus in a specific sibling’s genome, the probability that any given sibling inherited the same paternal haplotype is ½. Therefore, the probability that all *n* other siblings inherited the other paternal haplotype is 2^−*n*^. If any one other sibling did inherit the same paternal haplotype, the first sibling’s genome will be accessible for DNM calling at the locus we are considering. This implies that in our hypothetical family with one parent and *n*+1 children, a fraction 1 - 2^−*n*^ of all DNMs should be callable. For example, in Family 2, which consists of 3 children and their mother, ¾ of all DNMs should be callable.

Within each region that is accessible for DNM calling, most DNMs occurring on the proband’s maternal and paternal haplotypes should be identifiable, with the exception of DNMs that are shared with the sibling surrogate parent. Neglecting these sib-shared mutations, the mutation rate can be estimated by dividing the mutation count by two times the length of the accessible genomic region spanned by paternal IBD tracts. Moreover, read-backed phasing tools that are designed for use in parent/child trios can be applied with the surrogate father substituted for the father (Belyeu et al. 2021). Read-backed phasing will deduce that a mutation arose on a paternally inherited chromosome if it can be phased to a haplotype shared between the siblings that is not shared with their mother. Similarly, we can deduce that a mutation occurred on a maternally inherited chromosome if it can be phased to a haplotype that is shared with the maternal genome.

Since full siblings share DNA inherited from both of their parents, they can serve as surrogate parents for DNM calling even when the mother and father are both deceased, as is the case for the Generation I siblings P1, P2, P3, and P4 (for more details, see methods and **Figure 3A**). Since maternal and paternal chromosomes are passed down independently of one another, the fraction of DNMs accessible for calling in a family with *n*+1 children and no parents is expected to be (1 – 2^-*n*^)^2^ slightly smaller than the fraction 1 – 2^-*n*^ that is callable when paternal or maternal DNA is available. To call DNMs in P1, P2, P3, and P4 using surrogate parents alone, we first used hap-IBD to identify tracts shared identical by descent between pairs of siblings. For each sibling trio (P_i_, P_j_; P_k_), we then identified the set of regions where DNMs are callable in P_k_ using P_i_ and P_j_ as surrogate parents: this is the set of regions where P_k_ shares one haplotype IBD with P_i_ and shares the other haplotype IBD with P_j_ (**Figure 3B**). As long as two surrogate parents share distinct overlapping IBD tracts with the proband, we are able to randomly designate one as the surrogate mother and the other as the surrogate father and proceed with DNM calling without determining which haplotypes are actually maternally and paternally inherited (though we cannot use read-backed phasing to deduce whether mutations occurred on maternal versus paternal haplotypes). We also identified regions of the genome where one sibling was able to serve simultaneously as surrogate mother and father to another sibling; this is possible wherever the two siblings inherited the same haplotype from each of their parents, which occurs across about 25% of the genome in full siblings (**Figure 3C**).

**Figure 3.**
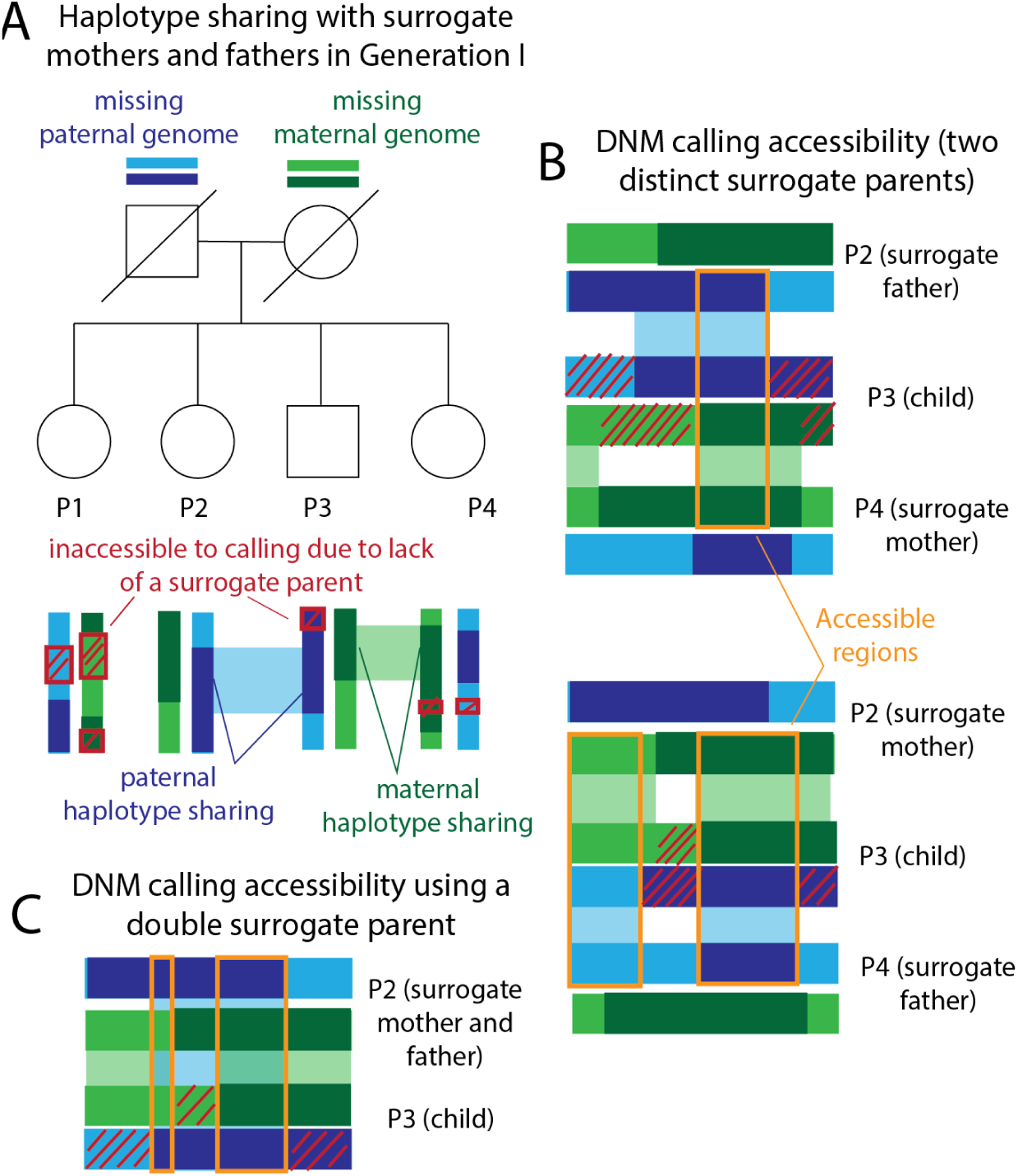
Calling DNMs using only surrogate parents. **A)** Generation I of our pedigree contains four full siblings (P1, P2, P3, P4) whose parents are deceased. We were able to call DNMs in these individuals using siblings as surrogate mothers and fathers. Example regions of paternal and maternal haplotype sharing are regions where P2 and P4 can act as surrogate father and mother to P3. Red crosshatches denote parental haplotype segments that were inherited by exactly one sibling–these regions are inaccessible to DNM calling due to lack of a surrogate parent. **B)** Illustration of maternal and paternal haplotype sharing in two example surrogate trios. DNMs are callable within orange regions due to haplotype sharing with both the surrogate mother and the surrogate father. **C)** Illustration of the regions that are accessible for calling in P3 using P2 as a double surrogate parent. This is possible when the surrogate and the proband share both maternal and paternal haplotypes.

To assess the quality of our surrogate-based DNM calls, we performed benchmarking using a family that we simulated as a composite of whole genome sequences from the 1000 Genomes project and mutation and recombination events from trios previously sequenced by Jónsson et. al (2017) (see Methods). We simulated a family with 5 total children, randomly selected one child as the “proband,” and called DNMs in this child using different subsets of the available real and surrogate parents. In both real and simulated families, DNM calling accessibility was similar to theoretical prediction (**Figure 4A**). As illustrated in **Figure 4B**, different sets of surrogate parents provided coverage of complementary genomic regions, and overlap between these tracts permitted error-correction of false positives that only appear when specific sets of surrogate parents are used. Overall inflation of the mutation rate by false positives is modest and appears to decrease with increasing family size (**Figure 4C-F**). Our simulation results suggest that caution is warranted when interpreting the mutation rates and spectra in Family 4, which contains just two siblings as well as their mother. Only half of the genome is accessible for DNM calling in this family (**Figure 4A, Figure S3**) and the ability to correct for paternal gene conversion will be limited, since none of the regions where DNMs are callable will contain both paternal haplotypes.

**Figure 4:**
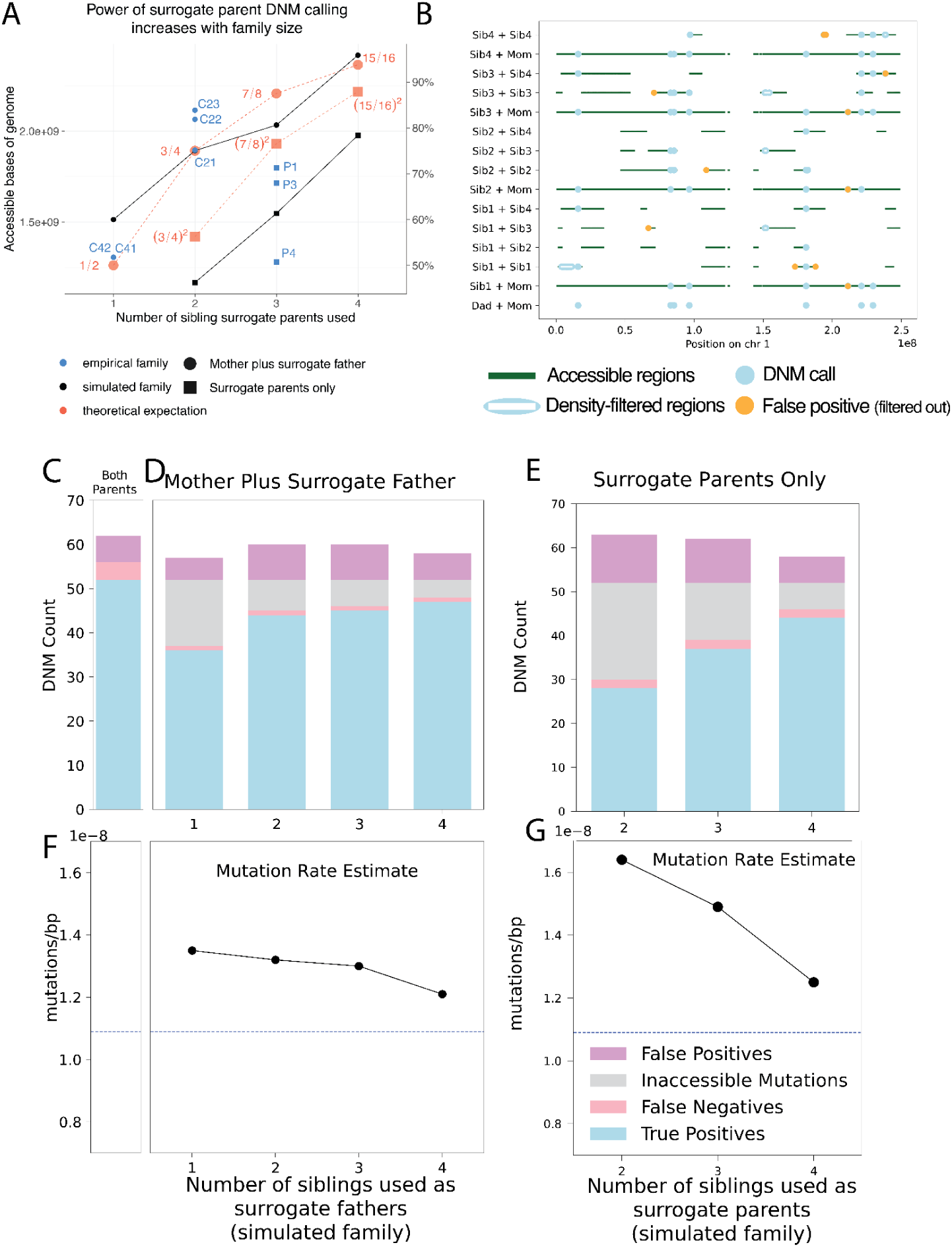
Precision and recall of surrogate-based DNM calling. **A)** The number of base pairs accessible for DNM calling in each individual from our pedigree (blue) depended on the number of real and surrogate parents available, broadly matching theoretical expectations (red dashed lines) as well as the proportion of accessible genome available for calling in our simulated pedigree (black) using similar configurations of real parents and siblings. Fractions in red indicate the proportion of the genome that is theoretically accessible. Families for which one real parent is available (circles) have a greater proportion of the genome accessible than those with no parents available (squares), but both improve when additional siblings are available. Theoretical expectations reflect the size of the genome accessible for SNP calling in the simulated family, which is close but not identical to the callable genome sizes in the empirical family. Empirical individual P2 was excluded due to somatic mutation contamination which lowered the accessible genome size (Figure S3; see discussion below). **B)** Genomic accessibility, DNM calls, and density-filtered regions in chromosome 1 of the simulated pedigree. Note that error correction is facilitated by overlap between regions accessible to calling using different sets of surrogate parents. **C)** True positive, false positive, and false negative DNM calls made in a simulated proband with both mother and father’s genomes available. **D)** As in C, but with a mother plus different numbers of siblings to act as surrogate fathers. Inaccessible mutations are DNMs that occur in regions of the genome without a suitable surrogate father. **E)** True positive, false positive, and false negative DNM calls made in a simulated proband with no parental genomes available, using different numbers of surrogate mothers and fathers. **F)** Mutation rates estimated with both simulated parents’ genomes available (dashed line). **G)** Mutation rates estimated using the mother’s genome and surrogate fathers. **H)** Mutation rates estimated using surrogate mothers and fathers, with no parental genomes available.

### Children of pathogenic MUTYH carriers have normal germline mutation rates

Previous studies have found that *MUTYH* variants specifically increase the C>A mutation rate in a variety of species and cell types (Sasani et al. 2022; Robinson et al. 2022). Since C>A comprises only about 10% of human DNMs, even relatively large perturbations of the C>A mutation rate are not necessarily enough to push the overall germline mutation rate significantly above its normal range, as previously seen in mice as well as humans (Sasani et al. 2022; Sherwood et al. 2023). In keeping with this expectation, we found most trios in this study to have normal mutation rates ranging from 8.23 × 10^-9^ to 2.14 × 10^-8^ mutations per base pair per generation (**Table S1**), comparable to the range between 7. 9 × 10^-9^ to 1.9 × 10^-8^expected in healthy individuals with parental ages between 15 and 50 based on a large previous study (Jónsson et al. 2017). However, we called a much higher frequency of mutations (4. 4 × 10^-8^mutations per site per generation) in the genome of P2, the biallelic mother of Family 2. Upon further examination, we found most of P2’s mutations to have unusually low variant allele frequencies (VAFs), between 20% and 50%. All other individuals had mutation VAF distributions centered around 50%, as expected of germline mutations that arose on one of two parental haplotypes (**Figure S4**). P2’s VAF skew suggests that most of their DNM calls are likely somatic mutations rather than germline mutations. Sherwood et al. (2023) previously noted a similar pattern in one of their biallelic *MUTYH* carriers who had undergone 5-fluorouracil chemotherapy for colorectal cancer, a treatment that can cause high-frequency mutations to emerge in the hematopoietic stem cell population. Due to this excess load of somatic mutations, which preclude estimation of an accurate germline mutation rate, we excluded P2 from further analysis and required a minimum VAF threshold of 30% for all mutations called in other individuals. Despite P2’s high somatic mutation rate, we found that their mutation spectrum was dominated by normal background mutational signatures SBS1 and SBS5, with no sign of the MUTYH-associated signatures SBS18/SBS36 (**Figure S5**).

### Testing the children of biallelic MUTYH carriers for skewed mutation spectra and parent-of-origin bias

Although we did not expect the overall mutation rate to be significantly elevated in trios with biallelic *MUTYH* carrier parents, we hypothesized that these families might have elevated proportions of C>A mutations and/or a higher-than-expected proportion of C>A mutations inherited from the affected parent. To maximize our power to test for these effects, we calculated trio-specific expected C>A mutation counts and proportions using a model fit to patterns of de novo mutations in 1,548 Icelandic trios with no known mutator phenotypes (Jónsson et al. 2017). Although this control dataset was generated separately from our study, we carried out similar filtering methodologies (**Figure S2A**), and all individuals in both studies are of European descent.

The Icelandic trio study by Jónsson et al. (2017) leveraged their data to predict the expected rate of each 1-mer mutation type per base pair per generation as a function of paternal and maternal age. Using this parental age model, we were able to calculate each trio’s expected maternal and paternal 1-mer mutation burden as a function of the parents’ ages at conception of the child (**Table S1**) and their accessible genome sizes (**Figure S6, Table S1**), following an approach recently used by Kaplanis, et al. (2022). For the most part, our empirical counts agreed with these expected counts (**Figure 5A**). In every child except for C42, the younger child with the abnormally high mutation rate in the family where we previously flagged DNM calling issues, the observed total mutation burden is within the upper one-tailed Poisson 95% confidence interval expected under the parental age model (**Figure S7**).

**Figure 5.**
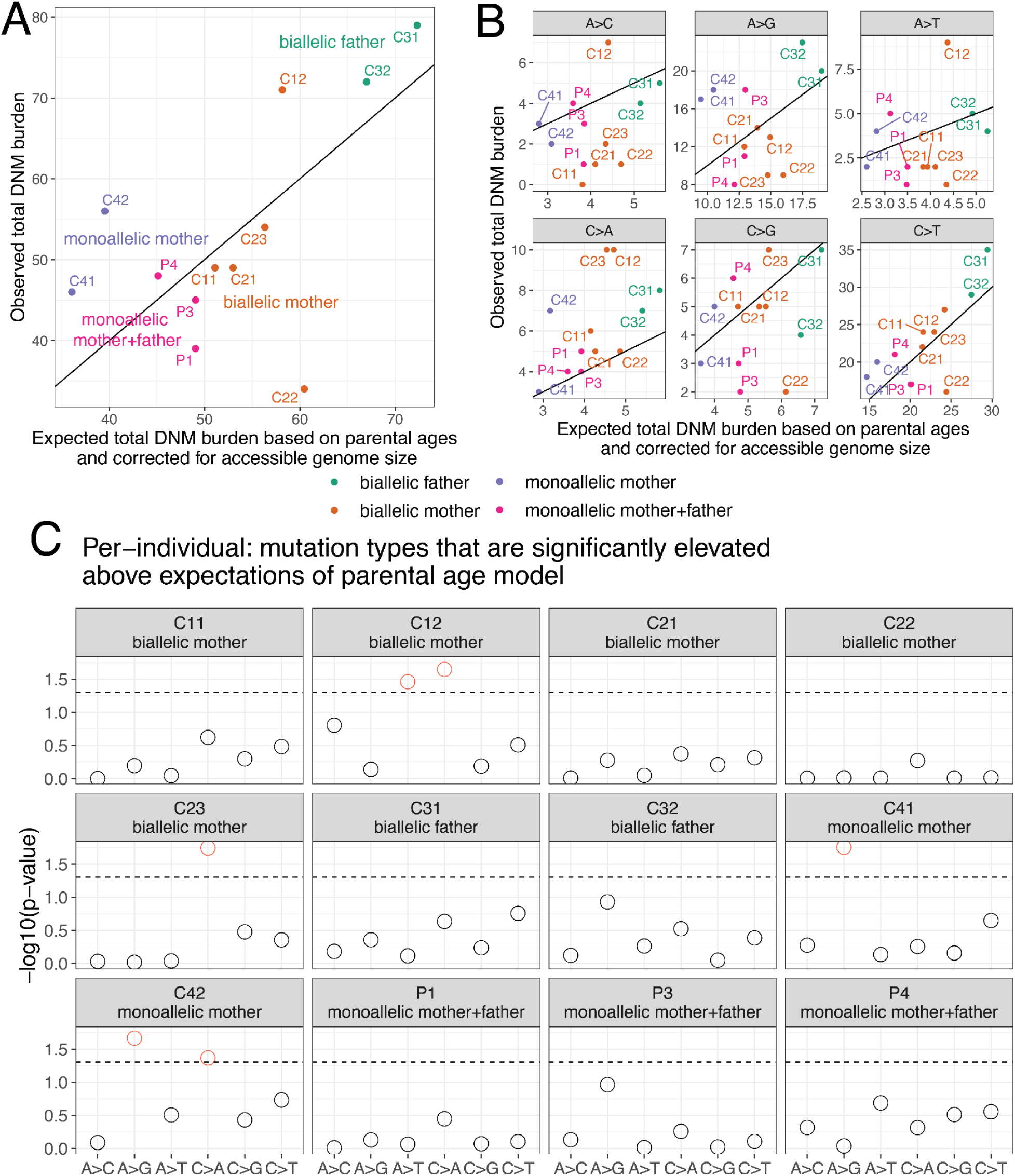
Observed and expected mutation counts. **A)** Comparison of observed DNM counts per child and the corresponding expected DNM counts under the parental age model (Jónsson et al. 2017), corrected for accessible genome size (**Figure S6**). P2 was excluded as discussed above due to evidence for somatic mutation contamination. Points are colored by the *MUTYH* carrier status of the child’s parent(s). Each child except for C42 has an overall mutation count that is compatible with the Jónsson parental age model (**Figure S7**). See **Figure S8** for a comparison with the results of Sherwood et al. (2023). **B)** Observed and expected mutation counts, faceted by 1-mer mutation type. Note that C>A counts are above the *y* = *x* line for nearly all individuals. **C)** The probability of observing a mutation count of each of the six 1-mer mutation types under the parental age model that is greater than or equal to what we observed for each member of the pedigree. Points above the dashed line (red circles) fall below the upper one-tailed Poisson *p* < 0.05 significance threshold. C12 and C23, both children of biallelic mothers, show significant elevation of C>A DNM counts, as does C42 (child of a monoallelic mother).

When we categorized DNMs by 1-mer mutation type (**Table S2**), we found that individual trio mutation spectra were largely consistent with the parental age model (**Figure S9**), but that C>A is the mutation type whose observed counts were most consistently inflated above expected counts, exceeding the expected count in all 12 individuals (**Figure 5B-5C**). Across the remaining five 1-mer mutation types, the proportion of trios exceeding the mutation count predicted by the parental age model ranged from 3/12 trios (A>C mutations) to 9/12 trios (C>T mutations) (**Figure 5B**). Most of the elevated C>A counts fell within an upper one-tailed 95% Poisson confidence interval of the expected count, but three children’s C>A burdens significantly exceeded the parental age model expectation (**Figure 5C**). These included C42 (one of our bioinformatic outliers), but also included C12 and C23, the children of two different biallelic mothers. The only non-C>A counts significantly exceeding the parental age model expectation were A>G mutations in C41 and C42 (a possible signal of inherited germline variant bleed-through due to the surrogate-calling method) and A>T mutations in C12 (**Figure 5C**).

We then added up sibling mutation counts to estimate the aggregate C>A enrichment within each nuclear family and found that the two families with biallelic mothers (Families 1 and 2) were enriched for C>A mutations by 1.81-fold and 1.46-fold above the expectation of the parental age model, respectively (**Figure 6A**). In Family 1, the total C>A mutation burden significantly exceeded the 95% upper 1-tailed confidence interval of the parental age model, while Family 2 falls 1 mutation below this significance threshold (**Figure 6B**). These C>A enrichments are comparable to the 1.57-fold-elevated C>A mutator phenotype recently identified in the mouse strain DBA/2J, but much less dramatic than the 6.04-fold enrichment phenotype identified in the mouse strain BXD68 caused by the homozygous loss of function R182Q-like mutation (**Figure 6A**). In contrast, neither child with a biallelic father had a significantly elevated C>A mutation load, and their family (Family 3) was only enriched 1.34-fold for C>A mutations overall (**Figure 6A-6B**). C>A mutation load was only 1.14-fold elevated (also nonsignificant) in the four parents P1–P4, whose own parents were all monoallelic and thus not expected to have a germline mutator phenotype. In Family 4, the family with a biallelic mother and significant bioinformatic obstacles to accurate DNM calling, we observed a non-significant 1.65-fold C>A enrichment along with a significant 1.75-fold A>G enrichment (**Figure 6A-6B**). A 1.65-fold C>A enrichment fails to reach significance in Family 4 because the siblings C41 and C42 each have a smaller callable genome proportion than individuals from families with a father or third sibling available for genotype calling. As in Sherwood et al. (2023), we then further summed up the mutation counts for children with the same carrier parent type, which in the case of our pedigree means summing up Families 1 and 2 which both have a biallelic mother as the carrier parent. This biallelic mother group showed significant enrichment of C>A DNMs (**Figure S10**).

**Figure 6.**
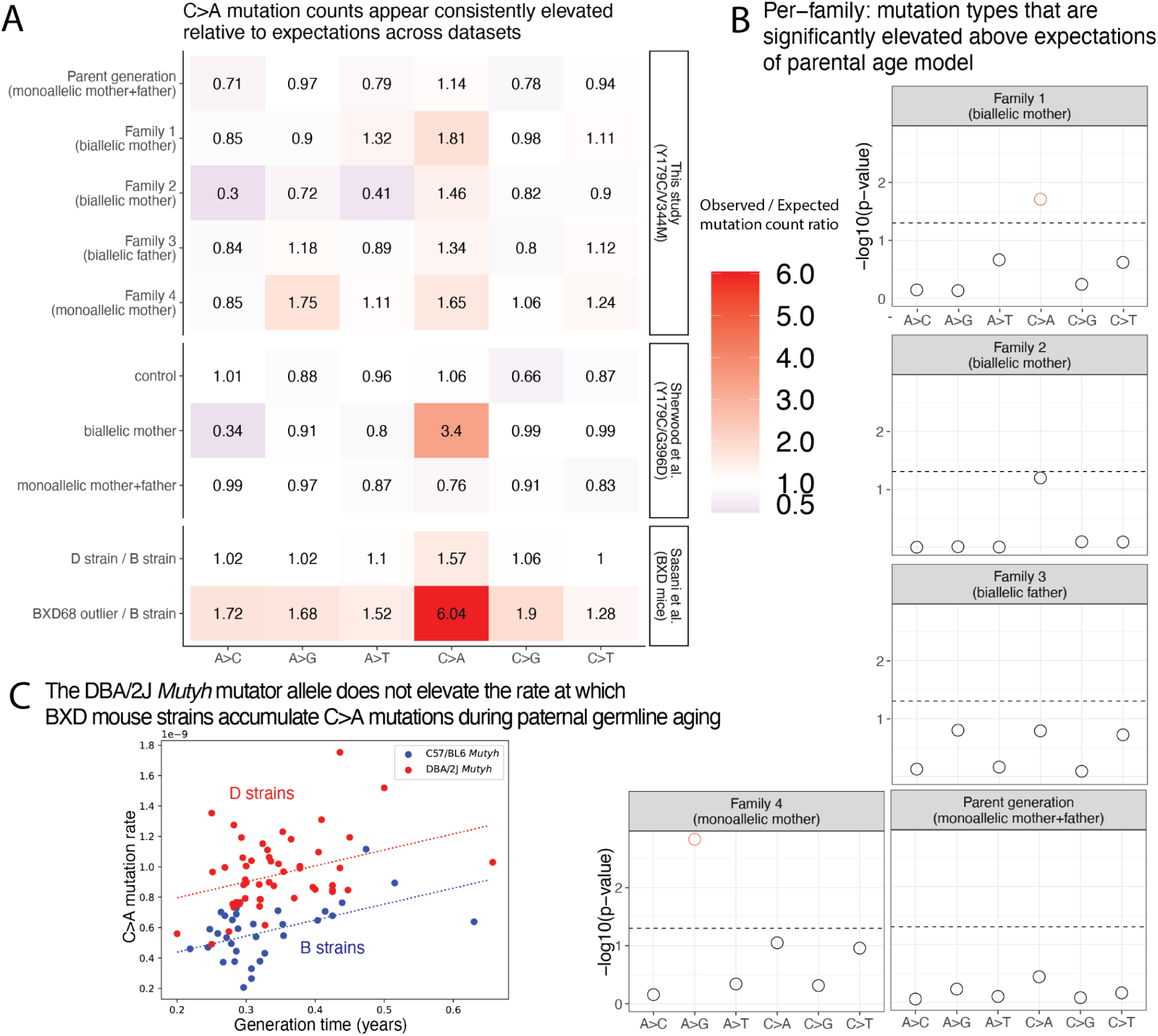
Children of mothers with biallelic *MUTYH* genotypes show significantly elevated C>A DNM counts. **A)** A heatmap showing the ratio of the observed / expected mutation counts per family (calculated by summing up the mutation counts per mutation type across all children within a family). These ratios are compared to the observed / expected ratio for the groups in Sherwood et al. (2023) (control group, individuals with a biallelic mother, and individuals with monoallelic parents), with expectations calculated using the parental age model. The bottom two rows show results from Sasani et al. (2022) for inbred BXD mouse strains: the “D” strain has an elevated mutation rate relative to the “B” strain, which has been linked to variation in *Mutyh*, and BXD68 is a mouse individual with an extreme outlier C>A mutator phenotype caused by a homozygous loss of function nonsynonymous mutation. The mouse ratios compare the per-generation rate of each mutation type between sets of inbred BXD mouse strains with different *Mutyh* genotypes. **B)** The probability of observing a mutation count of each of the six 1-mer mutation types under the parental age model that is greater than or equal to what we observed for each family in the pedigree. Points above the dashed line (red circles) fall below the upper one-tailed Poisson *p* < 0.05 significance threshold. Familys 1 shows significant elevation for C>A DNM counts above what is expected under the parental age model, and Family 4 shows significant elevation of A>G mutations.**C)** Multilinear regression of C>A mutation rate per generation as a function of generation time across BXD mouse strains. The *Mutyh* allele affects the regression intercept but not the slope, implying that mutator strains and non-mutator strains accumulate C>A mutations at the same rate during parental aging but accumulate these mutations at different rates during embryonic development.

We confirmed that our parental age model significance-testing framework was able to distill some of the main findings of Sherwood et al.’s (2023) study of germline mutator effects: in particular, the combined C>A burden of the children of the Sherwood et al. biallelic mother exceeded the 95% one-tailed confidence interval of the parental age model (**Figure S9**). In addition, all children of *POLE* and *POLD1* variant carriers in Sherwood et al. (genotypes which appear to have much more severe germline mutator effects than *MUTYH*) significantly exceeded the C>A and A>G mutation burdens predicted under the parental age model (**Figure S9**). We calculated a significant 3.4-fold enrichment of C>A mutations above the parental age model expectation in the family with a biallelic maternal *MUTYH* genotype sequenced by Sherwood et al. (**Figure 6A; Figure S11)**, suggesting that this family’s p.Y179C/G368D genotype may have a more severe mutator phenotype than the p.Y179C/V234M genotype affecting our pedigree.

Unlike Sherwood et al. (2023), we did not detect a significant increase in overall DNMs phased to the haplotype of the carrier parent (**Figures S12-S13, Table S1**). However, we did detect a significant elevation in C>A mutations phased to the maternal haplotype in Family 1 (one of the two families in our pedigree where the mother is the biallelic *MUTYH* variant carrier) (**Figure S14, Table S3**), indicating that there may be a carrier-parent-specific elevation of C>A mutations in this family. We note that this result is based on very low sample sizes of phased de novo mutations: 3 C>A mutations phased to the maternal haplotype in Family 1, compared to an expectation of 0.79 mutations, and so may be largely driven by stochasticity. In their biallelic families, Sherwood et al. were able to detect the activity of COSMIC mutational signature SBS18, a signature associated with defective *MUTYH* DNA repair (Alexandrov et al. 2020). However, mutational signature analysis of our DNM data did not identify any activity of either of the *MUTYH-*associated signatures SBS18 or SBS36. This likely reflects the small total sample size of C>A mutations in our data (**Figure S15**) and should not be interpreted as evidence of absence of SBS18/SBS36.

### Estimating C>A mutator effect sizes in the maternal and paternal germline

One consistent feature of human germline mutagenesis is that only about 25% of mutations appear to arise in the maternal lineage. In a family where the mother’s *MUTYH* genotype is pathogenic but the father’s genotype is normal, any elevation of the C>A mutation rate observed in the children likely arises due to either excess mutations that arose in the oocyte prior to conception or due to postzygotic mutations. Even if a child has inherited a normal *MUTYH* allele from their father, their postzygotic mutations may still be enriched for C>A if they arise prior to the maternal-zygotic transition, when the embryo first begins to express paternally inherited genes.

Given that almost 75% of germline mutations originate in the spermatocytes, the minimum maternal C>A enrichment required to explain our data is expected to be larger than the overall C>A enrichments recorded in **Figure 6**. To estimate the maternal germline C>A mutator effects that are required to explain the data, we started with the observed C>A mutation counts in Families 1 and 2 and subtracted the maternal and paternal C>A counts expected under the parental age model (**Figure S16A**). We then added each excess C>A count to the expected maternal C>A count and computed the proportional inflation of this value above the expected maternal C>A count. Using this logic, we calculated that the 1.46-fold to 1.81-fold overall C>A rate elevations observed in Families 1 and 2 (**Figure 6A, S16A**) imply maternal C>A mutation rate elevations of 4.2-fold and 2.7-fold, respectively (**Figure S16B**). The maternal effect implied by the Sherwood et al.’s (2023) 3.4-fold increase in overall C>A count is even larger: this value translates to a 10.2-fold elevation of the maternal C>A mutation rate (**Figure S16B**).

Although Family 3’s nonsignificant 1.34-fold C>A mutation rate elevation is not much lower than Family 2’s C>A enrichment, it implies a much lower paternal C>A rate elevation of only 1.45-fold (**Figure S16B**). This is very different from the 4.2-fold and 2.7-fold maternal C>A rate elevations that we infer to affect the two mothers who share the same biallelic *MUTYH* genotype. To investigate the likelihood that we were simply underpowered to detect a male germline mutator effect in Family 3, we calculated a “mutator detection threshold” for each family, which is the minimum number of extra C>A mutations required to produce a significant deviation from the parental age model (horizontal black bars in **Figure S16A**).

We then calculated how much this minimum number of extra C>A mutations should inflate the germline rate in the parent with the biallelic *MUTYH* genotype: this is the minimum fold-elevation of the biallelic parent’s C>A mutation rate that we have power to detect (horizontal black bars in **Figure S16B**). **Figure S16B** compares these minimum effect sizes to the effect sizes estimated using our empirical data (orange points). According to these calculations, we should have power to detect a paternal C>A mutator effect of 1.8-fold or greater, which is notably smaller than the maternal effects supported by the data, yet exceeds the level of paternal C>A enrichment that is supported by the data. We also carried this analysis on a per-individual level, rather than summed per family (**Figure S17**). Although this analysis is based on a limited sample size of individuals, it suggests that *MUTYH* variants may have a proportionally stronger effect on the maternal germline compared to the paternal germline.

### Mutyh variation does not increase the strength of the paternal age effect in the BXD mice

To further investigate the etiology of *MUTYH*’s germline mutator effect, we turned our attention from the human genotype p.Y179C/V234M to the mutator allele affecting the murine homolog *Mutyh* in the mouse strain DBA/2J. This mutator allele, which we call *Mutyh-D*, occurs in half of the recombinant inbred mouse strains known as the BXDs, which are each descended from crosses of DBA/2J with the standard lab strain C57/BL6. Each BXD strain has been inbred for tens or hundreds of generations and was previously whole-genome sequenced, which allowed the average mutation rate over the inbreeding period to be measured with high precision (Sasani et al. 2022). These rates revealed that the “D strains,” which have DBA/2J ancestry at *Mutyh*, have higher C>A mutation rates than the “B strains” which have C57/BL6 ancestry at this locus (Sasani et al. 2022). We were able to leverage these data to measure how the C>A mutation rate depends on parental age in the D strains as opposed to the B strains. Each BXD mouse strain has been inbred for a known number of generations spanning a known number of years, and we used these records to calculate each strain’s average generation time. We found that these rates ranged from 0.2 years to 0.63 years, spanning more than half of the mouse reproductive lifespan.

We fit a multilinear regression model to infer the dependence of the C>A mutation rate on *Mutyh* genotype (B or D) jointly with parental age, letting the *y* intercept of the model be the C>A mutation rate at the minimum parental age of 0.2 years (**Figure 6C**). We inferred a parental age effect of 1.05 × 10^-9^ additional C>A mutations per site per year (ANCOVA *p* < 0.001), but found no significant interaction between this parental age effect and *Mutyh* genotype (ANCOVA *p* > 0.82), indicating that the rate of C>A mutations occurring during gamete aging does not appear to differ between the B and D strains. In contrast, the baseline C>A mutation rate at age 0.2 years differs significantly between the B and D strains (4. 39 × 10^-10^ versus 7. 96 × 10^-10^ mutations per site per generation; ANCOVA *p* < 0.001). This suggests that the elevated C>A mutation rate associated with the DBA/2J *Mutyh* allele is driven primarily by early embryonic mutations, not mutations that occur in the paternal (or maternal) gametes.

## Discussion

We have investigated the germline mutation rate and spectrum within a large extended family affected by a *MUTYH* genotype, p.Y179C/V234M, consisting of a relatively common pathogenic variant plus a rarer variant with conflicting interpretations. This family’s history of colon cancer previously suggested that the p.Y179C/V234M genotype had a pathogenic effect, and we were able to use a cell-based *in vitro* functional assay to classify p.V234M as a partial loss of function variant. By calling de novo mutations in the children of two mothers with the p.Y179C/V234M genotype, we documented a modest but significant maternal mutator effect that appears weaker than the maternal germline mutator effect recently discovered in the children of two mothers with the more common MAP-associated genotype p.Y179C/G368D (Sherwood et al. 2023). A complementary analysis of parental age dependence of the BXD mouse *Mutyh* mutator effect confirms that the mutator is unlikely to act on the paternal germline and is most likely to increase the mutation rate during embryonic development, similarly to a maternal mutator allele that Stendahl, et al. (2023) recently discovered in rhesus macaques.

Even in a pedigree as large as the one we study here, DNM data sparsity limits the power to estimate precise mutator effect sizes. Based on prior knowledge about the biology of *MUTYH*, we expected to see excess germline C>A mutations in the children of biallelic carriers, and though our data appear to support this hypothesis, the observed C>A enrichments are likely not extreme enough to survive a stringent Bonferroni correction for the number of distinct tests performed throughout the manuscript, let alone an agnostic scan for mutators affecting other mutation types. We did not attempt to formulate a less conservative multiple test correction by estimating the number of truly independent tests being performed, which would have been challenging to do given the nested nature of testing both individuals and larger nuclear families for the same mutator effect. To give readers an accurate sense of data heterogeneity and noise, we perform more tests than the minimum number required, computing C>A enrichments individual by individual and observing nominally significant enrichments in only a few children (*p*<0.05 in a one-tailed test without multiple testing correction).

To our knowledge, this study is the first to call DNMs in the children of a father with a biallelic *MUTYH* genotype. Since about three-fourths of human variation arises in the paternal germline, we expected to have more power to measure a germline mutator effect in this family compared to families with maternal *MUTYH* variation. We were thus quite surprised that this father was the only biallelic parent whose children did not have a significantly elevated C>A mutation load, suggesting that *MUTYH* variation has a proportionally weaker effect on the paternal germline. This result should be interpreted with caution given our small sample sizes, but it could indicate that oxidative stress causes a smaller proportion of mutations in spermatocytes compared to oocytes or the developing embryo, or else that spermatocytes rely more on DNA repair pathways not involving *MUTYH*. Although it is possible that the one biallelic father in our dataset is an outlier, our mouse analysis also fails to detect an effect of *Mutyh* variation on the spermatocytes, which would be expected to increase the parental age dependence of the C>A mutation rate, and points to the embryo as the most likely site of the mutator effect.

Maternal biallelic *MUTYH* genotypes could plausibly increase the rate of germline C>A mutations during the first few embryonic cell divisions prior to the maternal-zygotic transition; maternal DNA repair machinery is responsible for repairing embryonic DNA damage prior to the activation of zygotic transcription (Huang et al. 2014; Harland et al. 2017). Early embryonic mutations are enriched for C>A, possibly due to 8-oxoguanine damage that occurs during oocyte and spermatocyte maturation, and maternal *MUTYH* mutations may interfere with the repair of such damage (Ohno et al. 2014; Smith et al. 2013b; Gao et al. 2019). Advanced maternal age appears to increase the rate of C>A mutations occurring on the paternal haplotype of the embryo (Gao et al. 2019), and biallelic maternal *MUTYH* mutations might cause a similar attenuation of 8-oxoguanine repair during the earliest stage of development.

One possibility is that 8-oxoguanine lesions cause similar absolute numbers of mutations per generation in males and females, but that the excess male mutation load is caused by factors unrelated to oxidative stress, which would seemingly contradict the widespread assertion that oxidative stress is a major cause of DNA damage in aging sperm (Aitken et al. 2003; Aitken 2020; Aitken and Krausz 2001). Further study of germline mutagenesis in families with paternal *MUTYH* mutations may thus shed light on the etiology of germline mutagenesis in males with normal *MUTYH* genotypes, helping us better understand whether oxidative stress is truly to blame for age-related infertility and the genetic disorders associated with paternal age. Our results suggest that 8-oxoguanine lesions may beget a larger fraction of oocyte mutations, making oxidative stress a notable contributor to reproductive decline in the general female population.

Because germline mutator phenotypes appear to be rare, at least at the current limits of our ability to detect them, these phenotypes have often been measured in the offspring of just one carrier parent, leaving us no information about whether these phenotypes are sex-specific. The mutator phenotypes recently measured by Kaplanis et al. (2022) were mostly found to affect male parents, and a study of an extended family affected by a DNA polymerase delta mutator definitively measured a stronger effect in male carriers compared to female carriers (Andrianova et al. 2023). Though the *MBD4* mutator allele that was recently discovered in rhesus macaques clearly exerts a maternal effect, the absence of a male breeder with the same phenotype precluded estimation of the relative strength of the corresponding male mutator phenotype (Stendahl et al. 2023). Pedigree studies like ours and the work of Andrianova et al. (2023) will likely be instrumental for further study of possible sex differences affecting mutagenesis and DNA repair.

A technical innovation that improved the power of this study was new methodology for calling DNMs in incomplete nuclear families, with siblings acting as surrogate parents. Given our goal of calling DNMs in the children of individuals with rare pathogenic *MUTYH* genotypes, we were able to maximize our pool of study subjects by relaxing the usual restriction to calling DNMs only in children whose parents’ genomes were both available for sequencing. Our surrogate parent approach does have drawbacks compared to DNM calling in complete nuclear families, most notably the restriction of the callable genome to regions where the proband inherited the same missing parental haplotypes as at least one sequenced sibling. Surrogate-based DNM calling is also susceptible to false positives caused by gene conversion and errors in IBD calling, and we note that these false positives will look cleanly mapped upon visual inspection of sequencing reads. The filters we employed to control these errors may have increased our false negative rates, particularly when using both a surrogate mother and a surrogate father. Conversely, our false positive rates appear to be elevated when only a single sibling is available as a surrogate parent and sib-sharing cannot be used to identify false positive DNMs that were actually inherited from the missing parent.

One new technology that will likely improve the performance of the surrogate method in the future is high-fidelity long read sequencing, which enables nearly all mutations to be phased to their maternal or paternal haplotype of origin (Noyes et al. 2022; Porubsky et al. 2024). When comparing the genomes of siblings who were sequenced using PacBio HiFi or a similar technology, it will become very straightforward to determine whether any read from the proband is derived from the same haplotype as a read present in a parent or surrogate parent, which will limit the ability of inherited variants to masquerade as DNMs. Surrogate-based mutation calling has the potential to make DNM analysis accessible to the many families where parents are deceased or not in contact with their children, including families affected by rare or undiagnosed genetic diseases where DNM calling is likely to have the greatest scientific and clinical utility.

Our data suggests that the germline mutator effect of *MUTYH* predominantly operates in a recessive manner, paralleling its role in cancer predisposition. However, we note that all available data on C>A mutation rates in normal human cells is derived from individuals who have at least one loss of function allele (p.Y179C). Although our study and previous studies (Sherwood et al. 2023) find mutagenesis and cancer risk to be associated with biallelic genotypes that combine p.Y179C with a partial loss-of-function allele (p.V234M or p.G368D), we do not have similar data from biallelic genotypes that combine two partial loss of function alleles, and we still lack an estimate of the human germline effect of two complete loss of function alleles. As we move toward better quantification of partial loss-of-function genotypes, it will be important to consider how they interact epistatically with each other and additional genes–for example, variants that impair the function of *MUTYH* and *OGG1* appear to interact epistatically in both the germline and the soma (Robinson et al. 2022; Sasani et al. 2023).

The apparent effect size difference between p.Y179C/V234M and p.Y179C/G368D suggests that there may be utility in moving beyond the binary classification of *MUTYH* variants as simply pathogenic or non-pathogenic. Although data sparsity issues imply that this effect size difference should be interpreted with caution, recent studies of *MUTYH* mutator alleles in the mouse germline and the human soma have also found that some genotypes have more severe mutator phenotypes than others. Previous somatic mutation data found an effect size difference between the common genotypes p.Y179C/G368D and p.Y179C/Y179C that appeared concordant with an earlier age of polyposis onset in p.Y179C/Y179C carriers (Robinson et al. 2022). For a rare genotype like p.Y179C/V234M, epidemiological data can likely not predict variant effect severity, and sequencing of normal tissues obtained from carriers of this genotype may prove to be a more viable option for obtaining this information. In this way, the mutation load in healthy tissues like the germline might eventually prove useful for predicting the severity of cancer risk likely to be associated with different pathogenic *MUTYH* genotypes, allowing clinicians to use whole genome sequencing to discern whether a family or an individual with a suspicious DNA repair variant is accumulating mutations in normal tissues faster than expected and might be at elevated risk of acquiring a mutation that transforms normal tissue into cancer.

## Data Availability

All data produced in the present study will be submitted to a secured human subjects data repository (dbGaP) so that other researchers may request access.

## Acknowledgements

We thank all the study participants for their time and engagement with our research. We thank Martha Horike-Pyne for her assistance drafting consent forms and applying for Institutional Review Board approval, and we thank Jailanie Kaganovsky and Vidha Sudhesh for their assistance mailing DNA collection kits to the study participants. We also thank the editor and three anonymous reviewers for providing feedback that helped us improve the manuscript. We also benefited from helpful discussions with Rosana Risques, Lea Starita, and members of the Harris Lab.

## Author Contributions

K.H. conceived the study. B.S. identified a suitable family for study, collected this family’s genotype and phenotype data, and led the design of the human subject engagement and biological sample collection protocol. K.H. contacted and consented the study participants. C.Y. coordinated the sample processing and genomic data generation. A.C.B., D.M.P., and K.H. designed the study’s computational analysis framework. Computational analyses were carried out by C.Y., A.C.B., D.M.P., K.H., and L.Z. The cell line variant assay was designed by J.O.K. and S.L.H. and executed by S.H. C.Y., A.C.B, K.H. and S.H. generated main text and SI figures. C.Y., A.C.B., and K.H. drafted the manuscript, and D.M.P., S.L.H., L.Z., J.O.K., and B.S. contributed manuscript edits.

## Data and Code Availability

All genomic data will be made available for controlled access via dbGaP. Per-individual de novo mutation counts and mutation spectra are available in Tables S1-S3. Custom scripts necessary for reproducing our analyses are available on GitHub at https://github.com/harrispopgen/mutyh_human_pedigree.

A human reference panel of phased VCF files from the high coverage 1000 Genomes project (Byrska-Bishop et al. 2022) was used to phase the data and infer shared haplotype tracts between relatives. These data can be found at https://www.internationalgenome.org/data-portal/data-collection/30x-grch38.

Poisson regression coefficients used for the parental age model can be found in Jónsson et al. (2017)’s Table S9. Sherwood et al. (2023)’s de novo mutation counts and mutation spectra are found in Table 1 and Table S2 of that study, respectively.

## Methods

### Recruitment and consenting of study subjects for biospecimen collection

The design of this study received prior approval from the University of Washington Institutional Review Board. After study participants gave written informed consent, they were each mailed an OGR-500 Oragene saliva collection kit. DNA was extracted from the Oragene kits at the Fred Hutchinson Cancer Center specimen processing core facility using the recommended standard protocol.

### Genome sequencing, SNP calling, and DNM calling

All sequencing was conducted at the University of Washington Northwest Genomics Center (NWGC). Samples had a detailed sample manifest (i.e., identification number/code, sex, DNA concentration, barcode, extraction method). Initial quality control (QC) entailed DNA quantification, sex typing, and molecular “fingerprinting” using a 63-SNP OpenArray assay derived from a custom exome SNP set. This “fingerprint” was used to identify potential sample handling errors and provided a unique genetic ID for each sample, which eliminated the possibility of sample assignment errors. Samples failed if: (1) the total amount, concentration, or integrity of DNA was too low; (2) the fingerprint assay produced poor genotype data; or (3) sex-typing was inconsistent with the sample manifest. No samples failed quality control at this stage.

Library construction was automated in 96-well plate format. At least 750 ng of genomic DNA was subjected to a series of library construction steps utilizing the KAPA Hyper Prep kit (KR0961 v1.14). All library construction steps were automated on the Perkin Elmer Janus platform. Libraries were validated using the Biorad CFX384 Real-Time System and KAPA Library Quantification Kit (KK4824). Barcoded genome libraries are pooled using liquid handling robotics prior to loading. Massively parallel sequencing-by-synthesis with fluorescently labeled, reversibly terminating nucleotides was carried out on the NovaSeq sequencer. Variant calling was carried out by the NWGC. Their variant calling pipeline combined a suite of Illumina software and other “industry standard” software packages (i.e., Genome Analysis ToolKit [GATK], Picard, BWA, SAMTools, and in-house custom scripts) and consisted of (1) alignment to human reference genome GRCh38DH using BWA-MEM (v0.7.15) (Li and Durbin 2009), (2) local realignment, (3) PCR duplicate removal (Picard MarkDuplicates; v2.6.0), (4) base quality score recalibration (BQSR) (GATK BaseRecalibrator; v3.7), (5) data merging, (6) variant detection, (7) genotyping, and (8) annotation.

Variant detection and genotyping were performed using the GATK HaplotypeCaller (4.2.0.0) (Van der Auwera 2020). Variants were initially flagged using the filtration walker (GATK) to mark sites that were of lower quality [e.g., low quality scores (Q50), allelic imbalance (ABHet 0.75), long homopolymer runs (HRun> 4) and/or low quality by depth (QD < 5)]. Data QC included an assessment of: (1) mean coverage; (2) fraction of genome covered greater than 10X; (3) duplicate rate; (4) mean insert size; (5) contamination ratio; (6) mean Q20 base coverage; (7) Transition/Transversion ratio (Ti/Tv); (8) fingerprint concordance > 99%; (9) sample homozygosity and heterozygosity; and (10) sample contamination validation. Genome completion was defined as having > 95% of the target read at > 10X coverage and > 90% of the target at > 20X coverage.

The SeattleSeq Annotation Server (http://gvs.gs.washington.edu/SeattleSeqAnnotation/), an automated pipeline, was used for annotation of variants derived from genome data. This publicly accessible server returned annotations including dbSNP rsID (or whether the coding variant was novel), gene names and accession numbers, predicted functional effect (e.g., splice-site, nonsynonymous, missense, etc.), protein positions and amino-acid changes, PolyPhen predictions, conservation scores (e.g., PhastCons, GERP), ancestral allele, dbSNP allele frequencies, and known clinical associations.

Putative de novo mutations in parent-offspring and surrogate-offspring trios were identified using the GATK(v4.2.6.1) PossibleDeNovo tool, which uses the genotype information from individuals in family trios to identify possible de novo mutations and the sample(s) in which they occur.

### Surrogate method

To identify haplotypes shared between relatives, we began by phasing the full 15-genome dataset using Beagle (Browning et al. 2021). In order to improve phasing quality, we phased these genomes together with a panel of 3,202 genomes from the high-coverage 1000 Genomes Project (Byrska-Bishop et al. 2022). Since rare variants are generally uninformative for identity-by-descent (IBD) segments, and are prone to sequencing error and phasing error, we filtered for common variants that are found at minor allele frequency > 10% in a subset of 2,504 genomes, and used them as the input to the program hap-IBD to infer shared tracts of IBD (Zhou et al. 2020). Additionally, the following hap-IBD parameter settings were used: min-seed=1.0, max-gap=1000, min-extend=0.2, min-output=2, min-markers=100. In this way, we were able to identify IBD segments that were shared between siblings but not present in any sequenced parent and then use these IBD segments as surrogates for missing paternal and maternal genome sequences. We noted that putative DNMs often occurred near the ends of our inferred surrogate parent tracts, and we hypothesized that these might be artifacts caused by inaccuracies in the boundaries of shared IBD tracts. To eliminate these artifacts, we implemented a density-based filter (see “Filtering” and **Figure S2B**).

We used GATK PossibleDeNovo as in the previous section on each informative “trio” of a child, a real parent if available, and one or two surrogate parents. For children whose parents’ genome sequences were both available (C11, C12, C31, and C32), we performed no surrogate DNM calling. For children whose mother’s genome was available but whose father’s genome was unavailable (C21, C22, C23, C41, C42), we called DNMs using the mother’s sequence plus each available relative as surrogate father. This resulted in two overlapping DNM call sets for each of the three siblings C21, C22, and C23, but just a single call set for C41 and C42. To generate each call set, we generated a positive mask file consisting of regions that we identified to be shared IBD between the child and the surrogate father, then called DNMs within the bounds of this positive mask minus the standard negative mask previously used to filter out low quality regions during standard DNM calling. We then merged together all call sets generated for the same child with different surrogate fathers.

To call mutations in each of the parents P1–P4, we ran PossibleDeNovo a total of nine times, each using a different combination of relatives as surrogate mother and father. Six of these runs involved a pair of two distinct relatives Pi and Pj, and the remaining three runs used the same sibling as both the surrogate mother and the surrogate father. For each run, a distinct positive mask was used to call mutations only in regions where the child shared two distinct parental haplotypes with its pair of surrogate parents. In the case where the same relative was used as both surrogate mother and surrogate father, this meant regions where the child shared two distinct IBD tracts with the same surrogate parent, because the two relatives had inherited the same chromosome from both their mother and their father. As before, DNM calls from all nine runs were merged to generate the total call set for each individual.

We generated additional mutation calls from P1–P4 by using each sibling P*i* as a “double surrogate parent” for each other sibling P*j*. We performed double surrogate calling within regions where hap-IBD found that P*i* and P*j* shared two overlapping IBD tracts, which indicates that they inherited the same maternal chromosome and also the same paternal chromosome. Since GATK PossibleDeNovo is designed for use with two distinct parental genomes, we called candidate double-surrogate DNMs within these double-IBD regions by identifying sites where the child is heterozygous but the double surrogate parent is homozygous.

All preliminary DNM calls were then filtered as described below (see “Filtering” and “IGV inspection.” During the IGV inspection step, we eliminated any putative DNM shared between two or more individuals, except when those individuals have a parent-child relationship, assuming that most such shared variants were in fact inherited from un-sequenced parents in regions erroneously identified as inherited IBD. In addition, we filtered out all DNMs called at sites that appear in the accessible regions of multiple surrogate parent combinations but are not consistently called using all of those surrogate parent combinations. For example, if a putative DNM in C22’s genome occurs at a locus that appears accessible for calling using either C21 or C23 as surrogate father, but that DNM is only called using C21 as surrogate father, it will be filtered out of the final call set. To eliminate additional false positive calls from the data, we manually removed any set of mutations that were clustered (over 6 mutations within 50 bp of one another) nearby a putative DNM with an rsID annotation, as we inferred these sites were more likely to have been inherited from a missing parent rather than a multi-nucleotide germline mutation event.

### Accessible Genome Size Estimation

Using both conventional Mendelian violation methods and our devised surrogate method, we derived the overall mutation rate for each offspring. Determining these rates required the computation of a denominator for each individual within the pedigree. This denominator represented the number of genomic sites where the read coverage was adequate (i.e., greater than 12 or less than 120) to ascertain a mutation, if present. Sites lacking confident inference of an individual’s parental haplotype sequences were excluded.

For offspring without sequenced fathers, our focus shifted to chromosomal regions where the child had an identical paternal haplotype with at least one sibling. For example, in the offspring of P2 with three children, two children with adequate read coverage at a site were necessary to identify mutations at that locus for both. For the parent generation, mutation identification depended on factors such as sufficient read coverage, successful haplotype reconstruction, and inheritance patterns. Using the surrogate method necessitated adjustments to the denominators based on the total length of shared parental haplotypes, leading to variable accessible base numbers for offspring in Families 2 and 4 and the parent generation (**Figure S3**).

### Filtering

DNMs were subjected to a series of quality control steps to eliminate potential false positives (**Figure S2A**). Building on prior research findings (Bergeron et al. 2022), true germline DNMs are usually characterized by alternative allele read support, with a variant allele frequency (VAF) ranging from 30% to 70%, and lack reads from either parent. DNMs were only considered for further analysis if they adhered to these parameters:

- Displayed a read depth between 12 and 120 for all members of both full pedigree and surrogate pedigree trios.
- Were identified by GATK PossibleDeNovo as being present in the child but not in either parent.
- Exhibited a VAF of 30-70% in the child.
- Had no reads supporting the variant in either parent.
- Genotypes filtered with GATK recommended hard filters: QD > 2.0; FS < 60.0; MQRankSum > -12.5; ReadPosRankSum > -8.0;SOR < 3.0

DNMs located in centromeres, telomeres, and segmental duplications were further excluded. Only DNMs that appeared in unique, accessible regions of the genome were retained in the final dataset. Additionally, any DNM that overlapped with variants having a minor allele frequency (MAF) of 1% or higher in the 1000 Genomes Phase 3 dataset was excluded. For DNMs identified using surrogate parents, a sliding window methodology was employed to pinpoint sparse mutations. The stipulated criteria for this was a maximum of 7 mutations within a 15MB sliding window, advancing in increments of 3MB.

### IGV inspection

In order to verify the mutation calls from both the full trio sequences and the resulting variants from families with surrogate parental sequences, we performed visual inspection of the resulting calls by inspecting the raw reads around the called de novo mutations.

We queried the original mapped sequences (bam files) to obtain all reads within 10kb (5kb slop) all pre-called de novo mutations in each trio of samples. When a mutation was detected in one of the families with a missing paternal genome we included all other samples in that trio that were used as a surrogate-paternal sequence, thus including multiple bam files as parental sequences.

The reduced files were then processed to filter low quality reads by selecting unduplicated sequences (-F 1024) and requiring a mapping quality higher than 20 (--min-MQ 20). To select informative reads used by GATK for variant calling, the unfiltered reads were also used to re-call variants using GATK HaplotypeCaller with the -bamout flag option that returns the informative reads for each call in bam format. The resulting variant files from this step were discarded and not used in any of the analysis. Note that if the algorithm would not return a mutation in that position there would be no informative reads available.

For each trio or surrogate-parent trio we generated a IGV report using igv-reports (github.com/igvteam/igv-reports) that outputs a HTML file containing small snippets of all called variants from the original vcf files. Each variant has 3 extra tracks per sample: (1) the original mapped sequence (bam file used in the mutation calling pipeline), (2) the filtered bams without duplicated or lower quality mapped reads, and (3) the bams of ‘informative reads’ yielded from the re-run of GATK HaplotypeCaller. These 3 tracks were included per sample in each trio, i.e. for a full trio a total of nine bam tracks will be included in the report while for a surrogate-parent trio the bams of all siblings and the available parents would be included. The reports included a 10Kb window around each variant and also included the allele count (AD, in each family) and the quality of the genotype (QD, in the original call).

Each variant in the IGV reports was then visually inspected to determine possible errors in the mutation dataset of each trio (**Figure S3**). The variants that failed our test were then classified according to their problematic features.

- Read evidence in the parental genomes, undetected due to indel realignment
- Read evidence in the parental genomes, undetected due to other reasons
- Unconventional or nuanced mapping
- Polymorphism evidence (as presence in dbSNP), for families with surrogate parents
- Polymorphism evidence (as presence in dbSNP), for families with surrogate parents
- A cluster of mutations (≥ 6 mutations per 50 bp), some or all of which have rsID annotations (indicative of misclassified germline mutations due to the surrogate-calling method)

This manual curation resulted in the number of DNMs being reduced by ∼36% per individual.

### Read-backed phasing

The tool Unfazed (v1.0.3) (Belyeu et al. 2021), a read-based phasing approach, was used to phase the de novo variants to maternal or paternal haplotypes. This approach required the existence of an “informative” inherited heterozygous variant that could be phased to a parent present on the same sequencing read as the DNM. This requirement resulted in 14-42% of DNMs being phased per individual (**Table S1**), a fraction typical for studies of phased de novo mutations.

### Generation and analysis of simulated trio data for benchmarking

To benchmark the performance of the surrogate method, we simulated a realistic pattern of mutation accumulation in a 5-sibling family and then simulated sequencing reads consistent with the family’s genotypes. In brief, we generated the family’s inherited germline variation using the 1,000 Genomes high coverage phased dataset (Byrska-Bishop et al. 2022) and then generated *de novo* mutations based on the data from Jónsson et al. (2017), both mapping to the hg38 assembly. Only autosomes were simulated.

More specifically, we sampled one “parent” at random from the 1000 Genomes CEU population and sampled the other at random from the GBR population, NA11893 and HG00132. This sampling was designed to match the European ancestry of our pedigree but avoid sampling individuals who were too closely related.

We randomly generated five “children” of these parents by generating recombination events using the recombination segments (*S_i_*) from a chromosome map (*M*) which was downloaded from the Beagle (2021) resource page (see https://bochet.gcc.biostat.washington.edu/beagle/genetic_maps/). We compiled the code to simulate the recombination events and combine the datasets from the multisample VCFs here, www.github.com/davidmasp/meiosim. Specifically, we simulated the number of crossings (*x*) using a Poisson distribution with λ equal to the length of the segment, measured in centimorgans (Δ*c*), multiplied by a recombination rate (*R*) of 0.01 (crossings/cM). We then obtained *x* crossings (*K_i_*) from a uniform distribution covering the positions of the recombination segment:

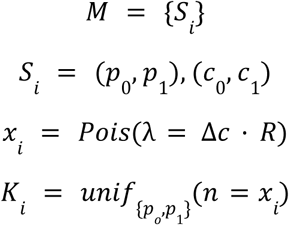

An initial haplotype was chosen at random from each parent and the haplotype was then swapped at each recombination breakpoint. SNPs from the parental haplotypes were then propagated to the children.

We added DNMs to each simulated child by selecting a proband uniformly at random without replacement from Jónsson et al. (2017) and editing the simulated child’s genome to include these DNMs.

We generated a short read BAM file consistent with each simulated genome using DWGSIM (0.1.15, www.github.com/nh13/DWGSIM), a tool that simulates sequencing reads from a reference genome and can incorporate custom germline variants. We defined an error rate of 0.001, a read length of 151 bp, and a target coverage of 30X. All other parameters were left as default. Other software used in this process were bcftools (1.19) and samtools (1.19).

We mapped and processed all simulated reads using SAREK (3.3.2) (Garcia et al. 2023). In brief, Sarek checks for quality and trims raw reads using fastqc (0.11.9) and fastp (0.23.4) (Chen et al. 2018); then maps with BWA-MEM1 (0.7.17-r1188) and further process them with Mark Duplicates and Base Quality Score Recalibration from the GATK suite (4.4.0.0) (McKenna et al. 2010). In addition to that, it measures the coverage using mosdepth (0.3.3) (Pedersen and Quinlan 2018) and integrates the quality reports with multiqc (1.15) (Ewels et al. 2016).

We then employed the same pipeline used for real families to process the jointly genotyped VCF file and simulated BAM files of the family. This involved applying the same filtering criteria for DNMs and utilizing identical genomic accessibility masks on the BAM files to calculate the final accessible genome denominators for each surrogate individual.

In simulating the surrogate method, we designated “sibling 0” as the child and performed DNM calling using all possible combinations of siblings 1-4 as surrogate parents, with scenarios including both the presence and absence of the real mother (HG00132) using GATK’s PossibleDeNovo. We then generated IBD tracks using hap-IBD to create surrogate tracts for all sibling pairs, in order to identify all regions where siblings shared a maternal or paternal IBD tract with sibling 0.

To minimize the inclusion of likely false positives, we applied the density filter (as detailed in the "Filtering" section and depicted in Fig. S2B) to retain only mutations in "sparse" genomic regions. While we did not curate these mutation calls via IGV, we did apply two stringent filters to remove false positive calls: 1) excluding any DNMs called in sibling 0 if another sibling had the alternate allele at that site, and 2) excluding any DNMs not called in all surrogate parent combinations which had the appropriate IBD segments to make that region of the genome accessible. We additionally classified false positive calls by identifying regions where a mutation was called in one surrogate trio combination but not in another, despite being accessible for detection. This distinction helped to account for limitations in the surrogate method and differentiate between true de novo mutations and likely false positives.

To assess how the inclusion of additional siblings as surrogates affected the method’s recall and precision, we conducted tests in two configurations:

- Group 1) one surrogate parent acting as the father with the real mother included, utilizing up to four siblings in various combinations (one sibling + mother, two siblings + mother, three siblings + mother, four siblings + mother)
- Group 2) two surrogate parents with no real parent, considering three potential combinations of sibling surrogates (two siblings used, three siblings used, four siblings used)

In each scenario, we subset the callset of true DNMs based on the accessible genome provided by these sibling combinations, thereby differentiating between false negatives and inaccessible mutations. We also applied a stepwise false positive filter by sites at which two or more siblings shared an alternate allele.

With each additional sibling included in the surrogate combinations, we assessed mutations that were accessible across multiple surrogate call sets but were called in fewer than expected, thus further reducing false positives. The mutation rate at each step was calculated similarly to the method described in the "Accessible Genome Size Estimation" section, with the number of true and false positives serving as the numerator and two times the size of the accessible genome as the denominator.

### Comparison to Jónsson model, ‘mouse model’, downstream statistics

Jónsson et al. (2017) carried out whole genome sequencing of Icelandic families and identified parental age impacts on the number and spectra of inherited de novo mutations. We used the Poisson regressions carried out in this study (listed in Table S9 of Jónsson et al. 2017) to predict expected de novo mutation burdens and spectra for each of the families in our study, based on parental ages.

In short, for each individual in our study, we plugged their parents’ paternal and maternal ages at the time of their birth into the following equations to get the expected count of each mutation type *c* (C>A, C>T, C>G, A>G, A>C, A>T):

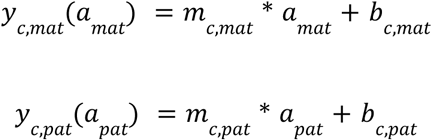

In these equations, *a*_*mat*_ and *a*_*pat*_are the maternal and paternal ages at the time of a child’s birth, respectively, *m*_*c*,*mat*_ and *m*_*c*,*pat*_ are the numbers of mutations of type *c* accumulated each year in the maternal and paternal germlines (linear regression slopes from Jónsson et al. (2017)’s Table S9), and *b*_*c*,*mat*_ and *b*_*c*,*pat*_ are the numbers of mutations of type *c* that would theoretically be present in the maternal and paternal germlines at age zero (mutation-type-specific maternal and paternal linear regression *y*-intercepts).

The resulting expected de novo mutation counts inherited from the mother and father add up to the expected burden of each type of de novo mutations in their child.

To correct for differences between Jónsson et al. (2017)’s accessible genome size (2.68 × 10^9^ bp) and the accessible genome sizes of each individual in our study (which ranged from 1.28 × 10^9^ bp to 2.67 × 10^9^ bp), we multiplied each expected mutation count under the parental age model by 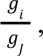 the ratio of the accessible genome of individual *i* (*g_i_*) to Jónsson et al. (2017)’s accessible genome size (*g_J_*). When the accessible genome size of an individual is considerably smaller than that of Jónsson et al. (as is the case for the individuals whose DNMs were called using the surrogate method), this rescaling will reduce the count of each mutation type we expect to observe in the offspring (**Figure S6**).

In order to determine whether the families in Sherwood et al. (2023) are consistent with the model trained on the families sequenced by Jónsson et al. (2017), we repeated the above procedure for the families in that study. Sherwood et al. (2023) didn’t report each individual’s accessible genome size, but since they did not employ the surrogate-calling method, their accessible genome size should be comparable to that of Jónsson et al. (2017), and so we did not carry out accessible genome size rescaling for these individuals. For each individual sequenced in our study and the Sherwood et al. (2023) study, we computed the ratio of observed to expected mutation counts for each mutation type.

When carrying out comparisons based on the subset of mutations we were able to phase to maternal and paternal haplotypes, we further downscaled the expected mutation counts by the phasing success rate per individual, which ranged from 14-40% (**Table S1**).

The above calculations yielded estimates of the relative rate of each mutation type in families with pathogenic human *MUTYH* genotypes relative to control families. To compare these effect sizes to the effect sizes of murine *Mutyh* mutator alleles, we computed analogous observed-over-expected ratios using mice with different *Mutyh* genotypes previously analyzed by Sasani et al. (2022). To compute the average mutation rate of each mutation type *c* in mice with a mutagenic *Mutyh* genotype known as the “D” genotype, we added up mutations of type *c* from all mice with the “D” genotype and divided this count by the total number of generations these mice were inbred, which is the total number of generations over which they had the opportunity to accumulate mutations. In the same way, we estimated a relative rate of mutations of type *c* in mice with the “B” *Mutyh* haplotype. Finally, we estimated the rate of mutations of type *c* in a single strain known as BXD68 affected by a unique *Mutyh* hypermutator phenotype. For the “D” allele and the BXD68 hypermutator allele, we divided the relative rate of each mutation type by the “B” allele rate to estimate the effect size of each of these *Mutyh* variants on mutagenesis in the mouse germline.

### Comparing our observed mutation counts to the null parental age model of Jónsson et al. (2017)

We used the Poisson cumulative distribution function (CDF) to determine whether the overall and per-mutation type DNM counts we observe are consistent with the parental age model, or whether we see significant elevations of any mutation type, particularly the C>A type associated with a defective MUTYH protein.

For each individual, we calculated *P*(*X* ≥ *k* | *λ*): the probability that a Poisson random variable *X* will generate a value greater than or equal to our observed mutation count *k*, given that it has mean *λ* equal to the expected count calculated based on the parental age model regressions from Jónsson et al. (2017) (as described above). We used *R*’s ppois() Poisson CDF function to calculate this probability. The ppois() function with the “lower.tail = F” flag gives the probability *P*(*X* > *k* | *λ*), and we calculated that *P*(*X* ≥ *k* | *λ*) = P(X > *k* -1 | *λ*), such that

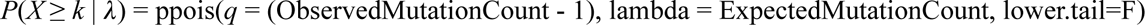

This approach was used to determine whether the total observed mutation counts per individual were significantly greater than what we’d expect under the null parental age model expectation. We separately carried out this analysis for each mutation type (C>A, C>G, C>T, A>G, A>T, A>C) per individual, per nuclear family, and for mutation counts phased to each parent (total counts and per-mutation type counts).

### Estimating the minimum mutator effect sizes that we have power to detect

For each biallelic parent whose offspring might be affected by a C>A mutator phenotype, we calculated the minimum C>A mutator effect size that should be statistically detectable using the above one-tailed Poisson test (leading us to reject the parental age model from Jónsson et al. 2017). To calculate this minimum effect size, we used the qpois() function in *R* to calculate the number of C>A mutations that should yield a p-value < 0.05, with *λ* estimated from the parental age model:

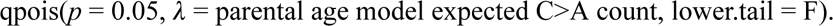

We then added +1 to the mutation count given by qpois() to calculate the number of mutations needed to be observed (*x*) such that P(*X* ≥ *x* | *λ*) < 0.05. We call this number of mutations the “mutator detection threshold.” We calculated separate thresholds for each child of a biallelic parent (including C11, C12, C21, C22, C23, C31, C32) and also calculated a cumulative threshold for detecting an elevated C>A mutation rate in each family with a biallelic parent (Families 1, 2 and 3). The detection threshold varies slightly across individuals and families based on parental age, the sex of the biallelic parent, and the childrens’ total accessible genome size.

To estimate the minimum biallelic *MUTYH* allele effect size we should be powered to detect, we assigned all excess C>A mutations above the parental age model’s expectations to the carrier parent:

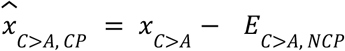

where *x*_*C*>*A*_ is the mutator detection threshold (minimum number of mutations for which P(*X* ≥ *x* |*λ*) < 0.05), *E*_*C*>*A*, *NCP*_ is the C>A count expected for the non-carrier parent (NCP) under the parental age model, and 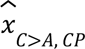 is the contribution of C>A mutations from the carrier parent (CP) needed to reach the significance threshold *x,* assuming all excess C>A above the parental age model expectation are assigned to the carrier parent.

The minimum detectable effect size of the biallelic *MUTYH* genotype should then be

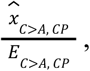

where *E*_*C*>*A*, *CP*_ is the expected number of C>A mutations contributed by the carrier parent under the parental age model.

We can also use this framework to estimate the effect size of the C>A mutator phenotype in the germline of each biallelic parent, again making the assumption that all excess C>A mutation counts above the parental age model expectation can be assigned to the carrier parent:

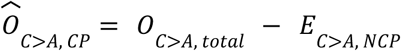

where *O*_*C*>*A*, *total*_ is the total observed C>A mutation count in an individual child or set of children of the same biallelic parent. As before, *E*_*C*>*A*, *NCP*_ is the expected number of C>A mutations contributed by the non-carrier parent under the parental age model, and 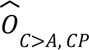 is the estimate of how many C>A mutations are contributed by the carrier parent, assuming all excess C>A mutations are assigned to that parent.

The *MUTYH* effect size required to yield this number of mutations is then

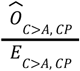

where *E*_*C*>*A*, *CP*_ is the expected number of C>A mutations contributed by the carrier parent under the parental age model.

### Mutational Signature Analysis

Non-negative matrix (NMF) factorization was used to extract mutational signatures from the de novo 3-mer mutation spectra, either per-individual, or summed up per-family. *SigProfilerExtractorR* (v. 1.1.16), an R wrapper for *SigProfilerExtractor* (Islam et al. 2022), was used to carry out the analyses. The reference genome was set to “GRCh38” and 100 NMF replicates were used. A range of signature numbers were explored, ranging from 1-10 for the per-individual analysis, and 1-3 for the per-family analysis (above 3 there were too many signatures for the number of input samples when individuals were grouped per family). The optimal solution that maximizes stability while minimizing cosine similarity was chosen by the software: for each analysis (per-individual and per-family), one signature was chosen as the optimal solution.

The cosine similarity between the optimal reconstructed mutation spectra and the empirical data ranged from 0.563--0.821 in the per-individual analysis, from 0.803-0.882 in the per-family analysis.

The optimal single signature in each analysis was deconvoluted by SigProfilerExtractor into contributions from known COSMIC (Catalogue of Somatic Mutations in Cancer) signatures. In each case, the extracted signature was deconvoluted into signatures SBS1 and SBS5, two clock-like signatures that generally make up the bulk of mutations in both germline and somatic data. No contributions of SBS18 or SBS36, somatic mutational signatures associated with defective *MUTYH*, were detected.

### Cellular assay of MUTYH glycosylase function

Human HEK293 *MUTYH* KO cell lines were transduced with lentivirus containing *MUTYH* cDNAs, either WT or variant, each cloned into pCW57.1 (Addgene #41393; gift from Dr. David Root). Transduced cells were selected and stable MUTYH expression was induced as previously described (Jia et al. 2021). To measure *MUTYH* variant function, cells expressing each variant were then co-transfected with a GFP reporter containing an 8oxoG:A mispair (Raetz et al. 2012; Nagel et al. 2014) and an mCherry-expressing plasmid as a transfection control. After a ∼72 hr incubation with the reporter, cells were analyzed via FACS with a BioRad Ze5. A function score was calculated as the fraction of repair positive (mCherry+, GFP+) cells out of all transfected cells (mCherry+), divided by the same quantity for cells transduced with WT MUTYH, and scaled by a log2 transform, such that a score of 0 indicates WT-like repair function, and negative scores indicate deficient function.

### Analysis of parental age dependence of the BXD mouse mutation rate

We downloaded data on the mutation rate per generation, B versus D haplotype status at the *Mutyh* locus, and the number of generations of inbreeding for each of the BXD mouse strains from the Github page associated with Sasani, et al. (2022) (https://github.com/tomsasani/bxd_mutator_manuscript). We then computed each strain’s C>A mutation rate per site per generation (number of years of inbreeding divided by number of generations of inbreeding) and fit an ordinary least squares multilinear regression to explain this variable as a function of generation time minus the minimum observed generation time of 0.2 as well as the categorical *Mutyh* status variable.

