## Supplemental Figures for "A maternal germline mutator phenotype in a family affected by heritable colorectal cancer"

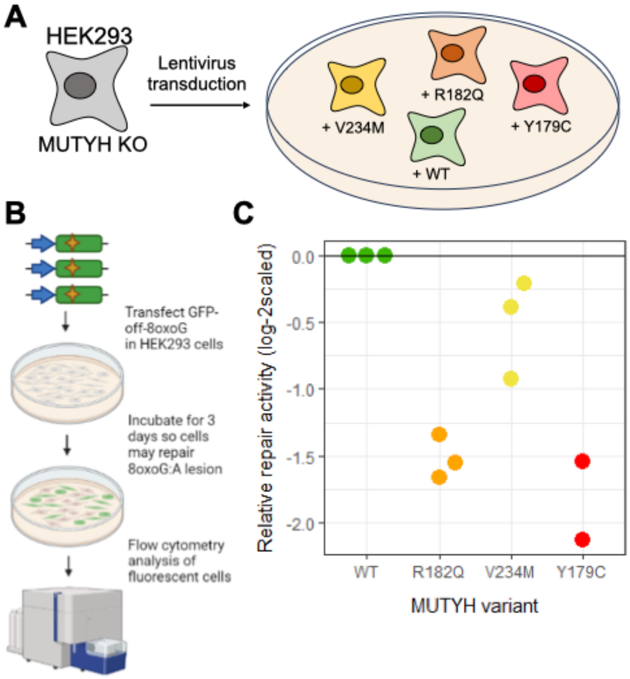


**Figure S1**. **Cell-based *in vitro* assay of MUTYH 8-oxoG:A repair function. A)** We first generated knock-in HEK293 cells expressing different MUTYH variants. **B**) We then transfected in a GFP reporter containing an 8-oxoG:A mispair, which turns cells green when the A is replaced with a C. Flow cytometry was used to sort cells based on GFP fluorescence. (Panel was generated using biorender.com) **C**) Results of the GFP-off assay, with the relative repair activity measured as the frequency of each variant in the GFP+ fraction compared to the frequency before sorting.

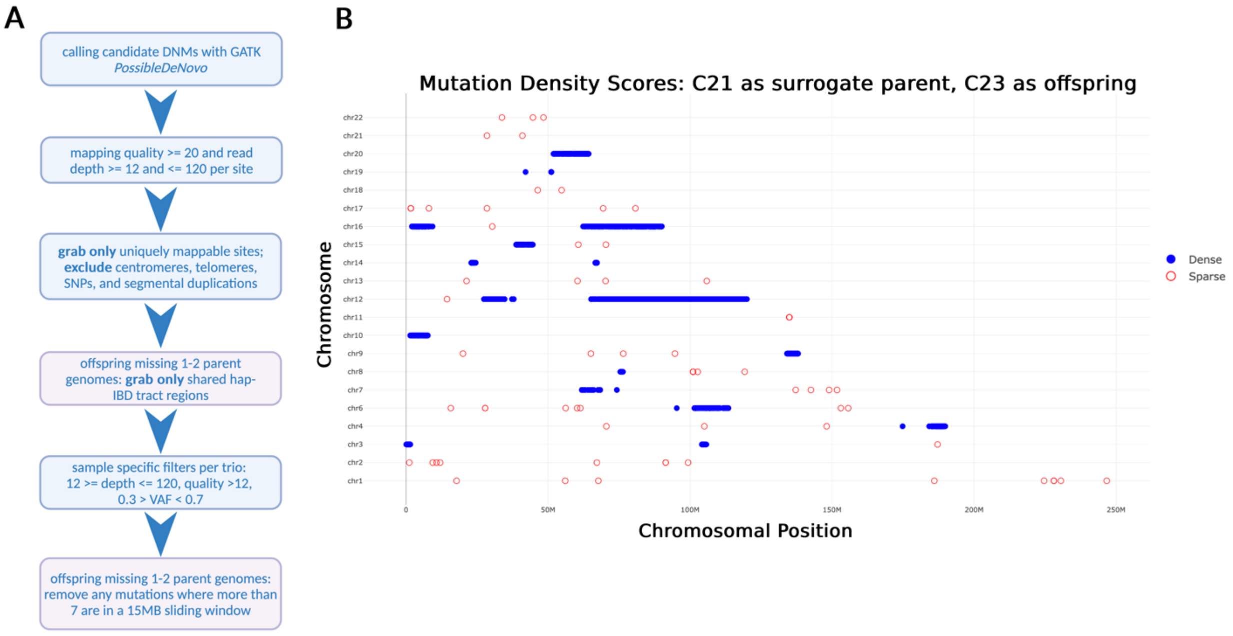


**Figure S2. DNM calling and assessment. A)** DNM calling and filtering workflow diagram. **B)** An example of the density filtering method used in the sibling-as-surrogate-parent calling method applied to C22 as the offspring of surrogate parent C21. Candidate DNMs are "sparse," meaning they are in regions where no more than 7 mutations were identified in sliding window sizes of 15MB (with step size of 3MB). Sparse mutations are outlined in red, while "dense" mutations (that did not pass this density filter) are outlined in blue, and likely represent regions of the genome where the two siblings did not share the same paternal haplotype.

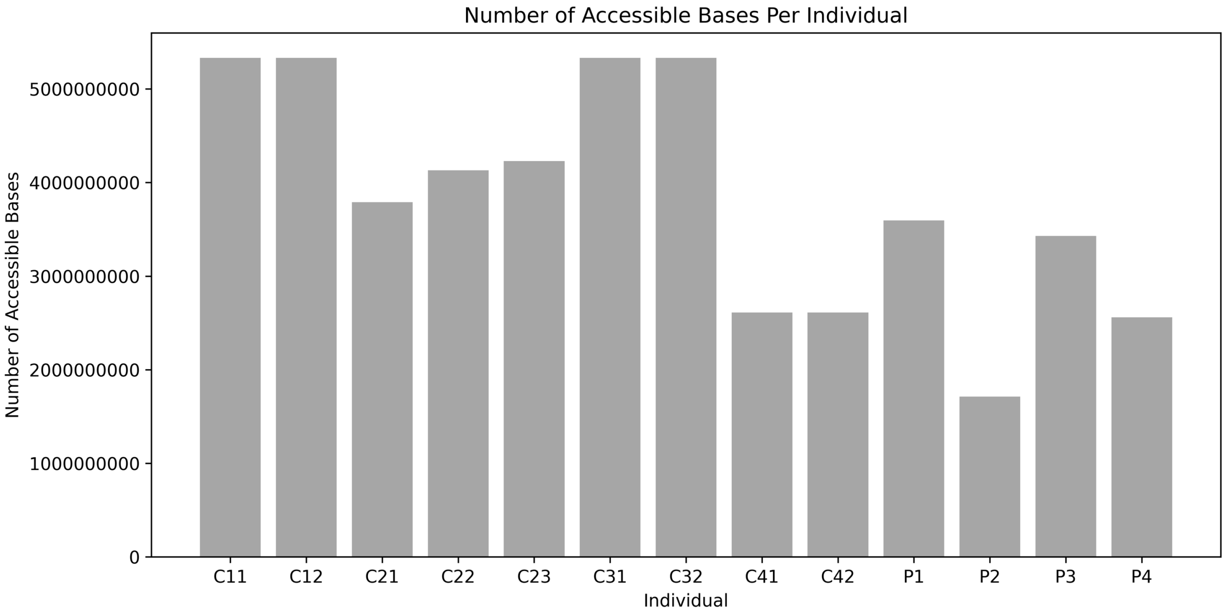


**Figure S3. Number of Accessible Bases Calculated per Individual.** The number of accessible bases identified per each sequenced individual (excluding S1 and S3). Individuals that did not require the surrogate DNM calling approach (C11, C12, C31, C32) share the same maximum number of accessible bases. Individuals in Families 2 and 4 and the parent generation all have a lower amount of accessible bases, which is dependent in each case on the number of shared paternal haplotype bases seen in each surrogate parent sibling combination. As there is only one combination for offspring of Family 4, the number of accessible bases is lowest in these individuals.

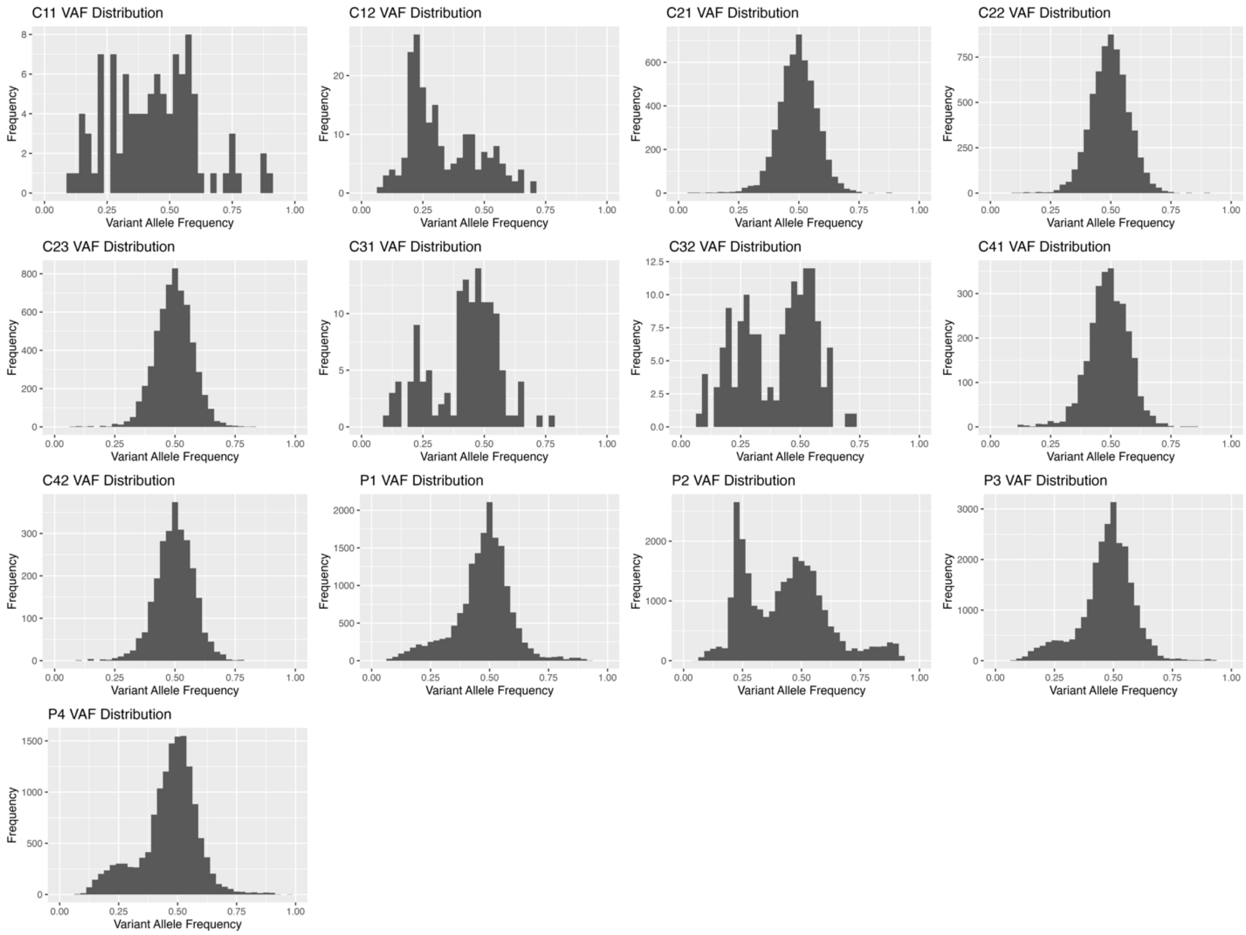


**Figure S4. VAF distributions of all candidate DNMs per individual.** Distributions of variant allele frequency (VAF) scores are displayed for all identified DNMs of the 13 studied individuals. Two modes are frequently observed across individuals, typically centering below and above 0.40- these likely reflect clonal somatic mutations and germline mutations, respectively. Note that P2 is a clear outlier, with a VAF distribution heavily skewed to the left with relatively few mutations in the candidate germline part of the distribution.

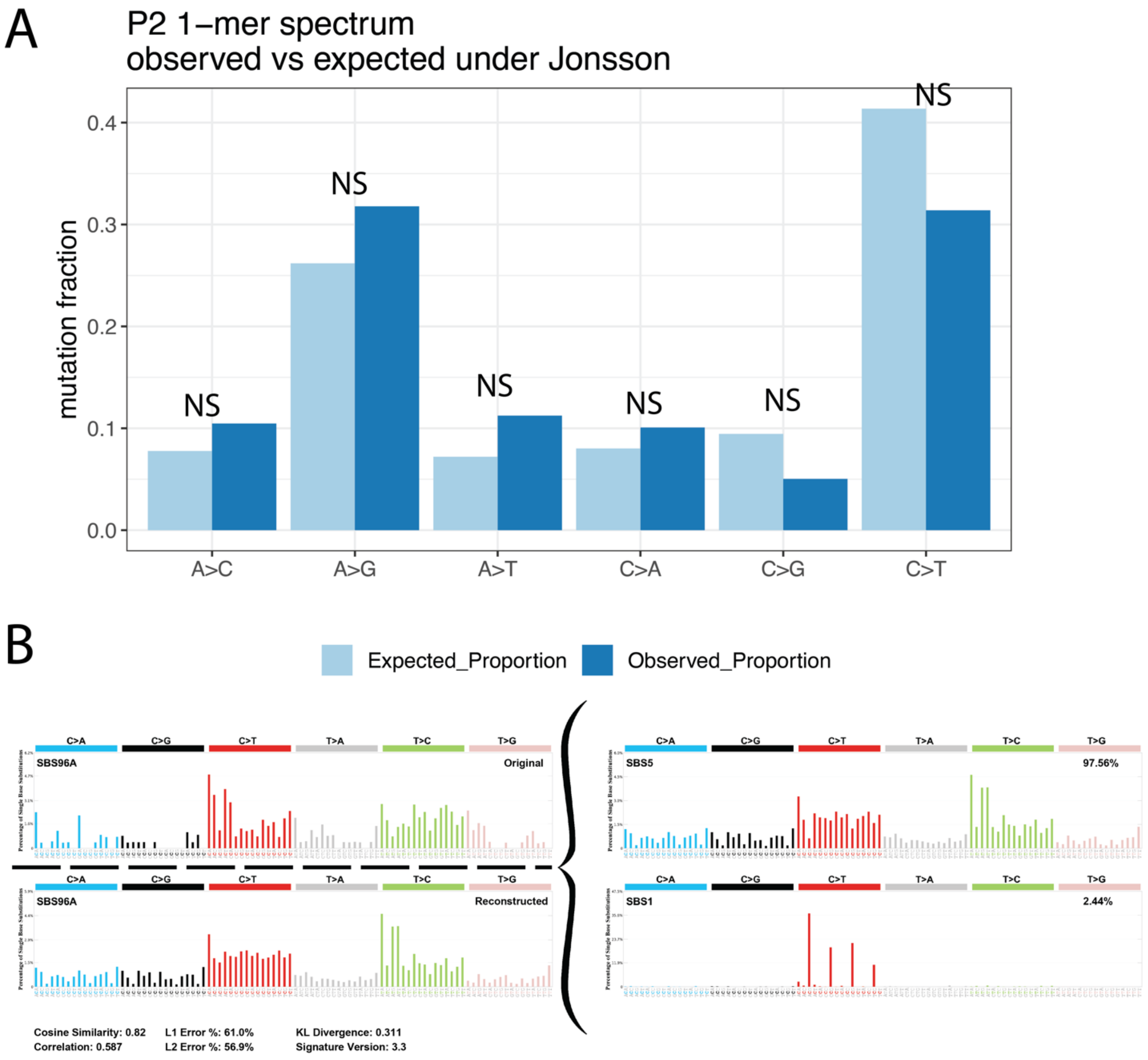


**Figure S5. Analysis of spectra in the individual (P2) with elevated number of mutations. A)** Comparison of the 1-mer de novo mutation spectrum for individual P2, who showed signs of somatic mutation contamination. The proportional 1-mer mutation spectrum was compared to that expected based on the ages of P2's parents at the time of their birth under the parental aging model. Each mutation type's count relative to the sum of the other five types was compared to model expectations using a 2x2 Chi-Squared test and Fisher's Exact test. No comparison resulted in a p-value < 0.05 (labeled as NS for non-significant), indicating that the spectrum is not significantly different from expectation. Note that these mutations did not go through extensive filtering or manual inspection due to P2 being excluded from further analysis early in the pipeline. The Chi-Squared test was used instead of the Poisson-based test for other individuals because P2's mutation counts were so highly elevated above the counts expected under the parental aging model, so comparing relative proportions of mutation types was more suitable. **B)** We carried out mutation signature decomposition on the 3-mer spectrum of P2 using SigProfilerExtractor and both the COSMIC v3.3 and v2 catalogs (results using v3.3 shown). The mutation signature was decomposed into SBS signatures 1 and 5; signatures SBS18 and SBS36 did not appear.

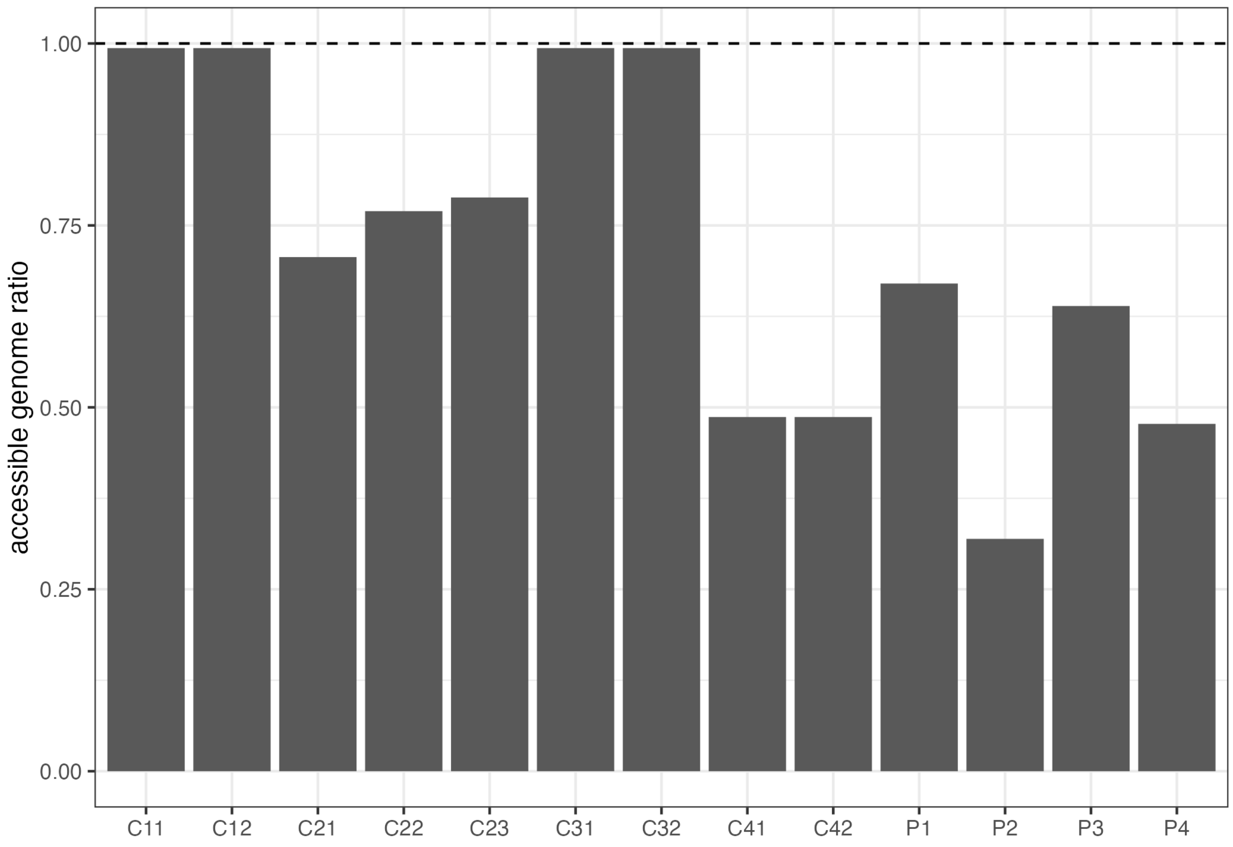


**Figure S6.** Ratio of accessible genome size for the individuals in this study over the average accessible genome size reported in Jonsson et al. (2017) (2,682,890,000 base pairs). Individuals C21, C22, C23, C41, C42, P1, P2, P3, and P4 all have accessible genome ratios substantially <1 due to the use of the surrogate-parent DNM calling method.

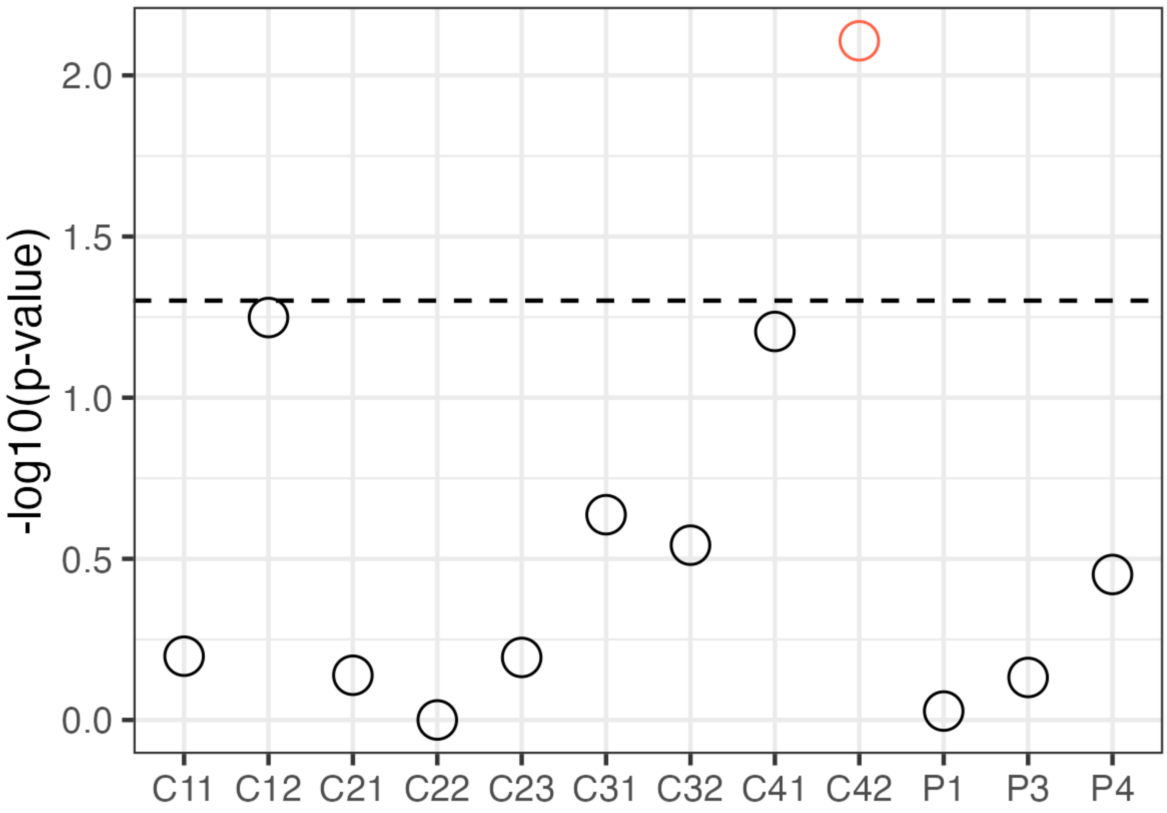


**Figure S7.** The probability (from the Poisson cumulative distribution) under the parental age model (Jonsson et al. 2017) of observing an overall mutation count greater than or equal to what we observe. The dashed line indicates the p-value threshold of 0.05 (significant points colored in red). All individuals other than C42 have overall DNM counts that are compatible with the parental age model. See the **Methods** for more detail on how probabilities are calculated.

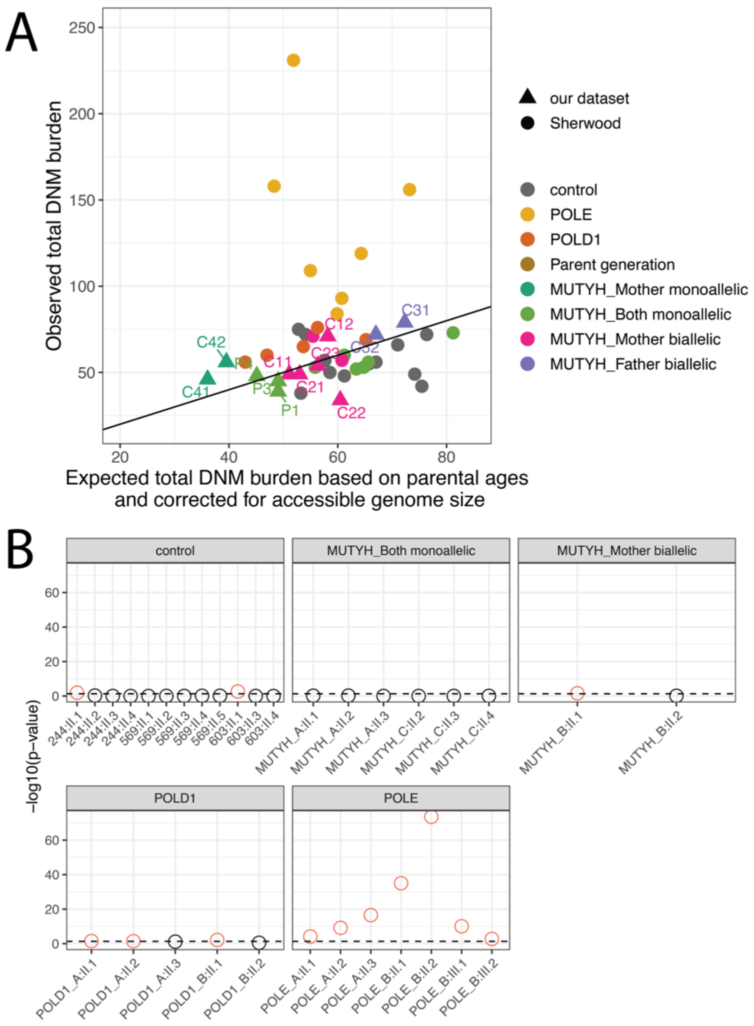


**Figure S8. Deviations from the parental age model in Sherwood et al. (2023). A)** Comparing observed and expected DNM burdens from individuals in this study (triangles) and individuals from Sherwood et al. (2023) (circles). Expected DNM burdens are based on parental age and accessible genome size for the individuals in this study, and based on parental age only for Sherwood et al. (2023), since accessible genome size was not reported (and was therefore assumed to be ~equivalent to the accessible genome size used to generate the parental age model in Jonsson et al.). "*POLE*" and "*POLD1*" refer to individuals in Sherwood et al.'s dataset that have variation in those polymerase genes and show an extreme effect on the germline mutation rate. The $y = x$ line is shown in black. As in Sherwood et al., individual *MUTYH*_C:II.1 was excluded due to high levels of somatic variant bleed-through. **B)** The probability (from the Poisson cumulative distribution) under the parental age model of observing an overall mutation count greater than or equal to what Sherwood et al. observed. The dashed line indicates the p-value threshold of 0.05 (significant points colored in red). Most of Sherwood et al.'s control individuals (except for two) have overall mutation counts consistent with the parental age model, as do all their individuals with monoallelic *MUTYH* parents. However, one of their individuals with a biallelic *MUTYH* mother has a significantly elevated DNM burden, and the majority of individuals with the more severe variants in *POLE* and *POLD1* show extremely significant elevations of overall mutation count.

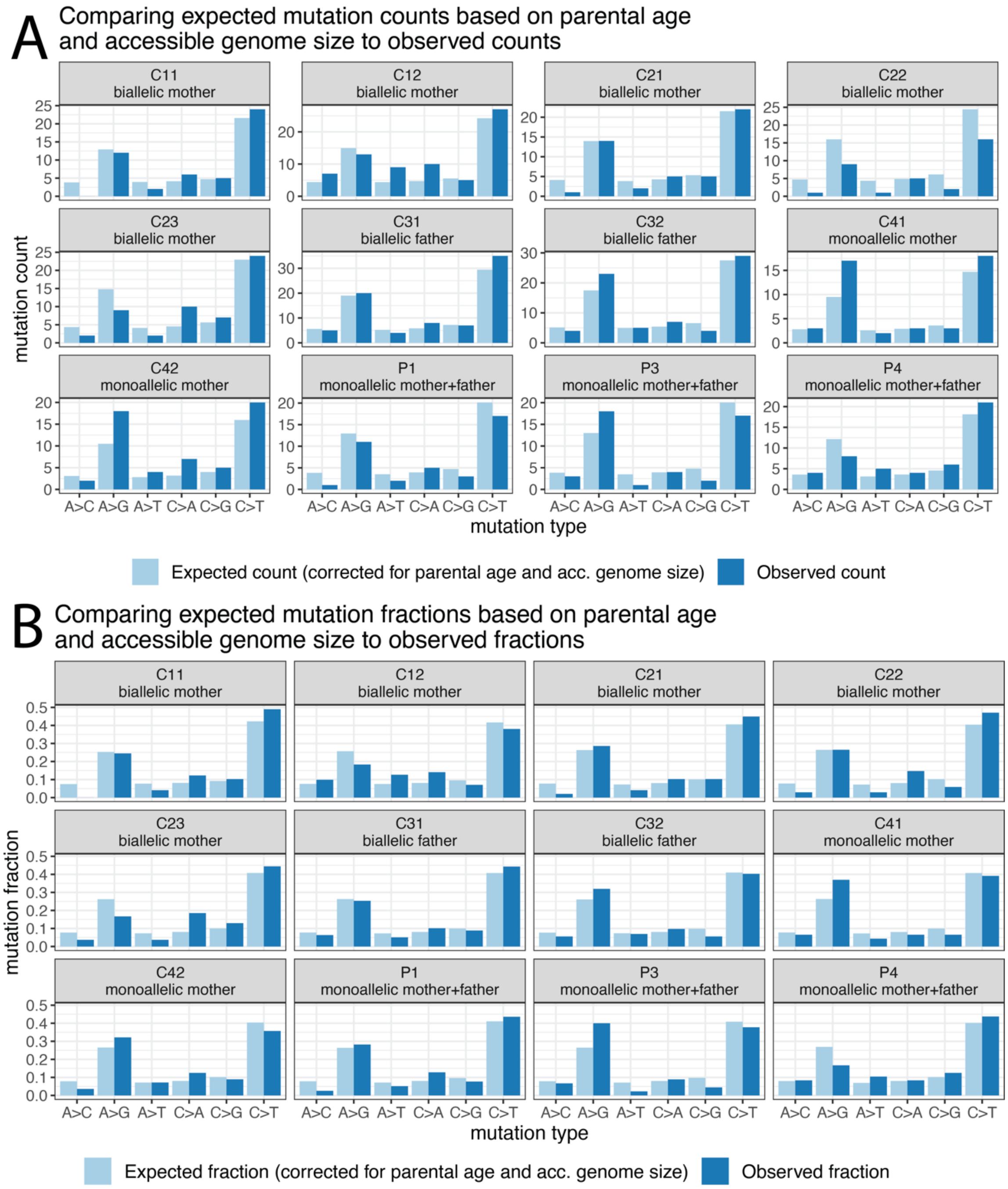


**Figure S9. Comparing observed and expected (under the parental age model) mutation spectra. A)** Comparing observed (dark blue) mutation counts for each 1-mer mutation type to expectations (light blue) under the parental age model. **B)** Comparing observed and expected mutation fractions (proportion of the total mutations) across mutation types.

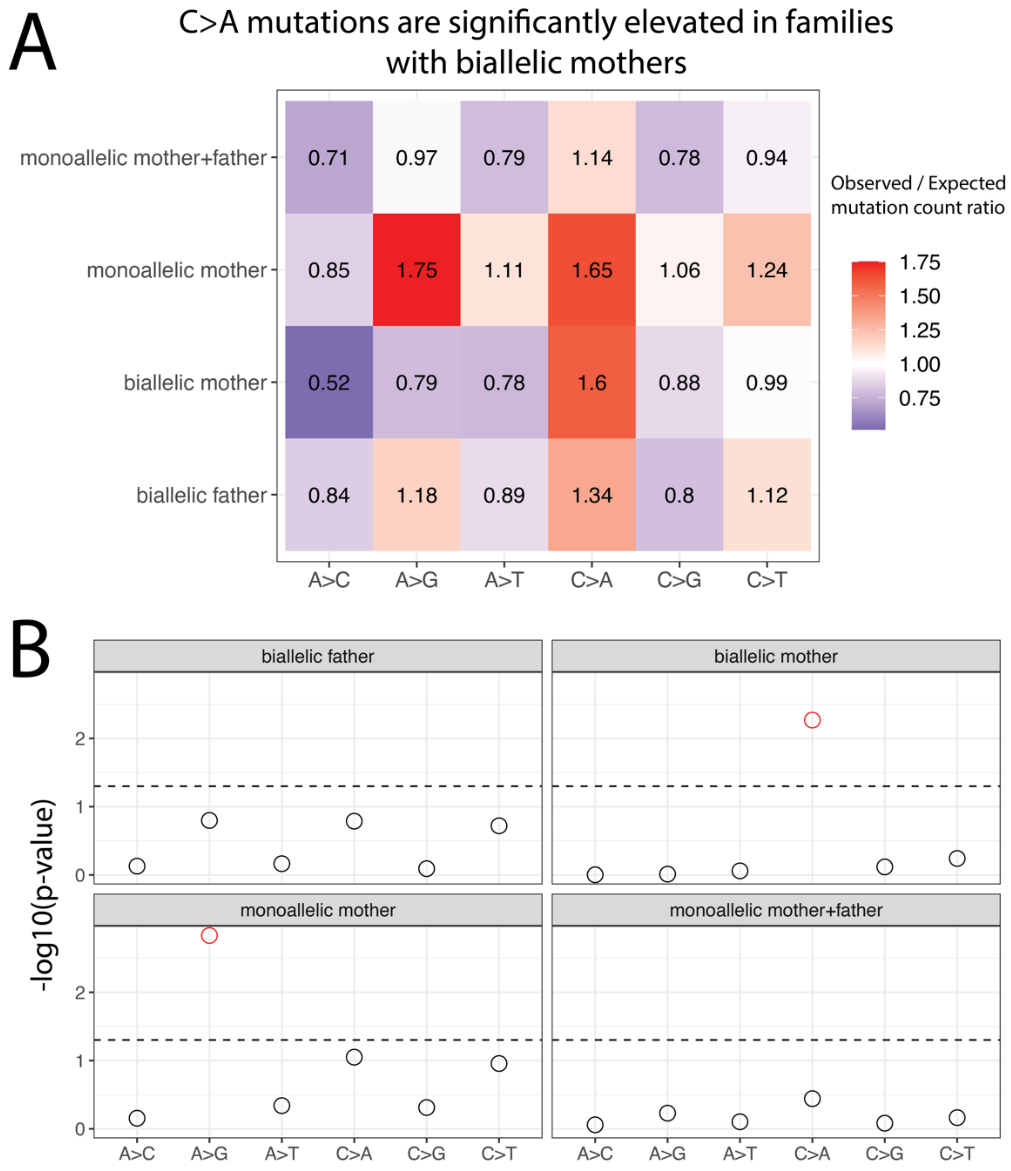


**Figure S10. Families with biallelic mothers show significant C>A elevation. A)** A heatmap showing the ratio of the observed / expected mutation counts per group of children in our dataset that have parents in the category listed on the y-axis (calculated by summing up the mutation counts per mutation type across all children with a parent of the types listed on the y-axis, as in Sherwood et al.). Only our ‘biallelic mother’ category contains more than one family (Families 1 and 2). The ‘monoallelic mother+father’ represents the parent generation individuals, the ‘monoallelic mother’ group is Family 4, and the biallelic father group is Family 3. **B)** The probability of observing a mutation count of each of the six 1-mer mutation types under the parental age model that is greater than or equal to what we observed for each group. Points above the dashed line (red circles) fall below the upper one-tailed Poisson $p < 0.05$ significance threshold. The biallelic mother group (Families 1 and 2) shows significant elevation for C>A DNM counts above what is expected under the parental age model, and the monoallelic mother group (Family 4) shows significant elevation of A>G mutation types above expectations.

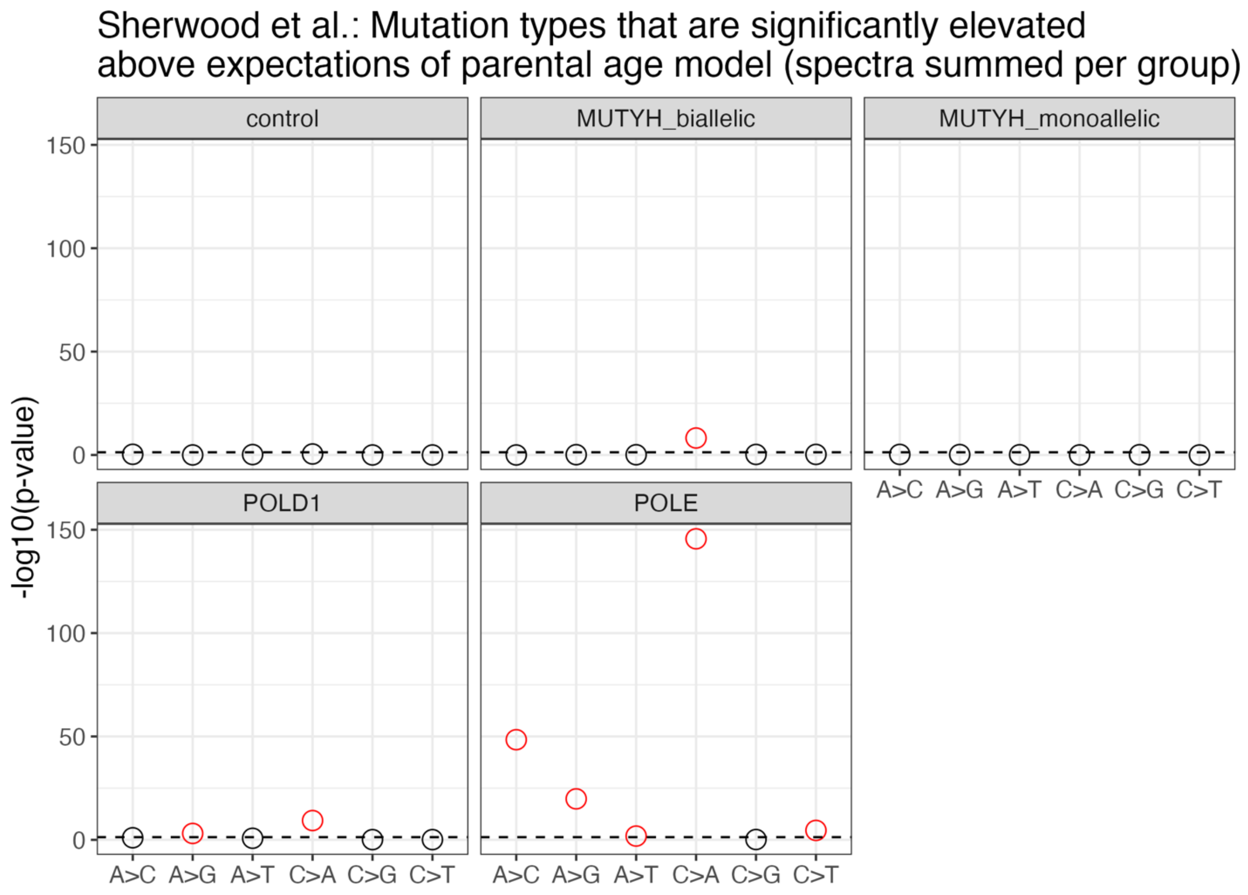


**Figure S11. Elevated C>A counts in biallelic *MUTYH*, *POLD1* and *POLE* groups from Sherwood et al. (2023).** Under our significance testing framework, the mutation spectra summed per group from Sherwood et al. (2023) (summarized in the heatmap in **Figure 4A**) show a significantly elevated C>A count for the biallelic *MUTYH* family, as well as the groups of individuals with more severe *POLD1* and *POLE* variants.

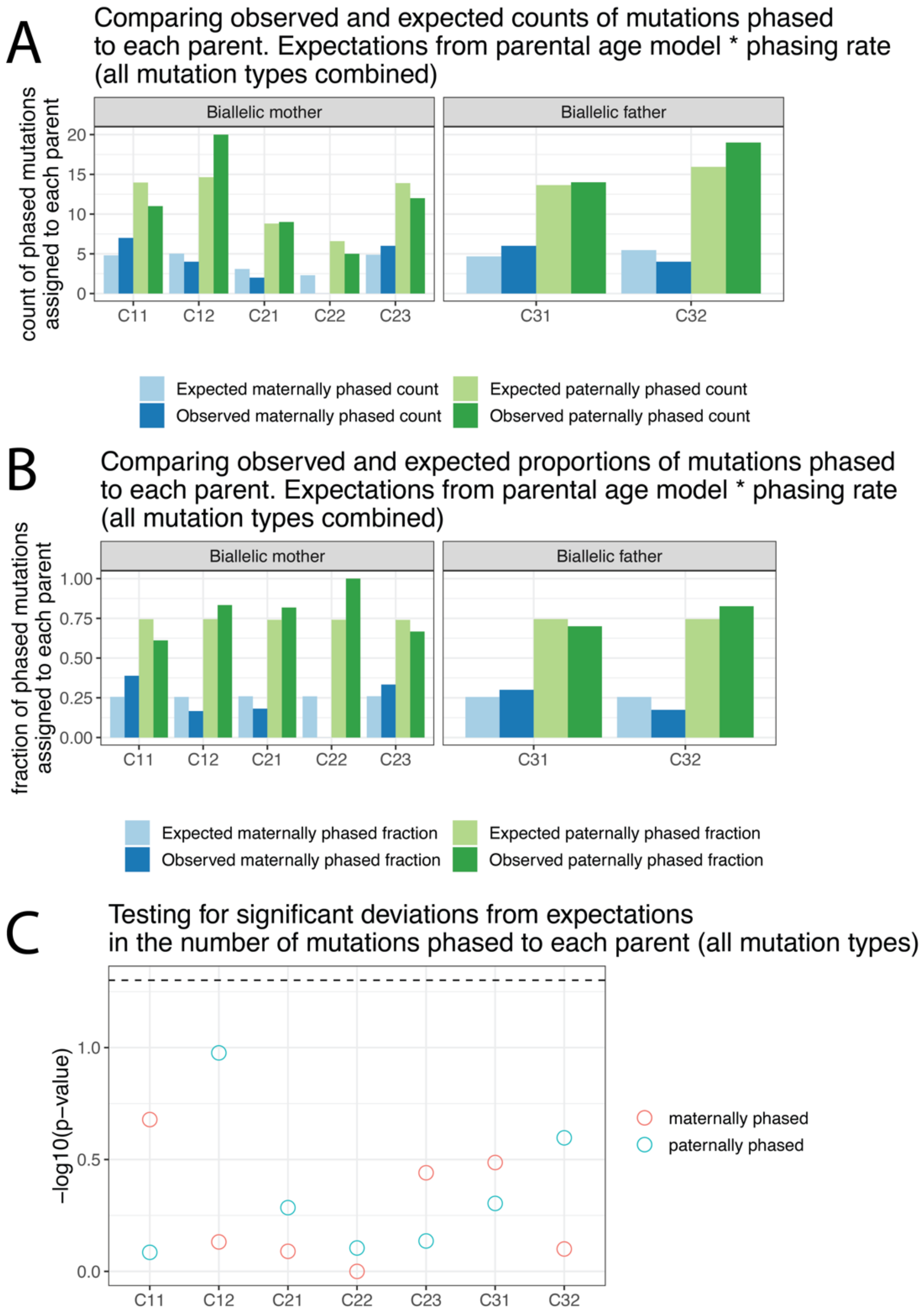


**Figure S12. No significant differences in the amount of mutations phased to the carrier parents are observed.** Counts **(A)** and relative fractions **(B)** of phased mutations assigned to either the maternal (dark blue) or paternal (dark green) haplotypes. Expectations (light blue and light green) are based on the number of mutations expected to come from each parent under the parental aging model (corrected for accessible genome size and individual phasing success rate). **C)** The probability (from the Poisson cumulative distribution) of observing greater than or equal to the number of mutations phased to each parent under the parental age model (corrected for accessible genome size and individual phasing success rate). Dashed line indicates $p < 0.05$ threshold. No individual shows significantly more mutations phased to either parent than in expectation. See the **Methods** for more details on how probabilities are calculated.

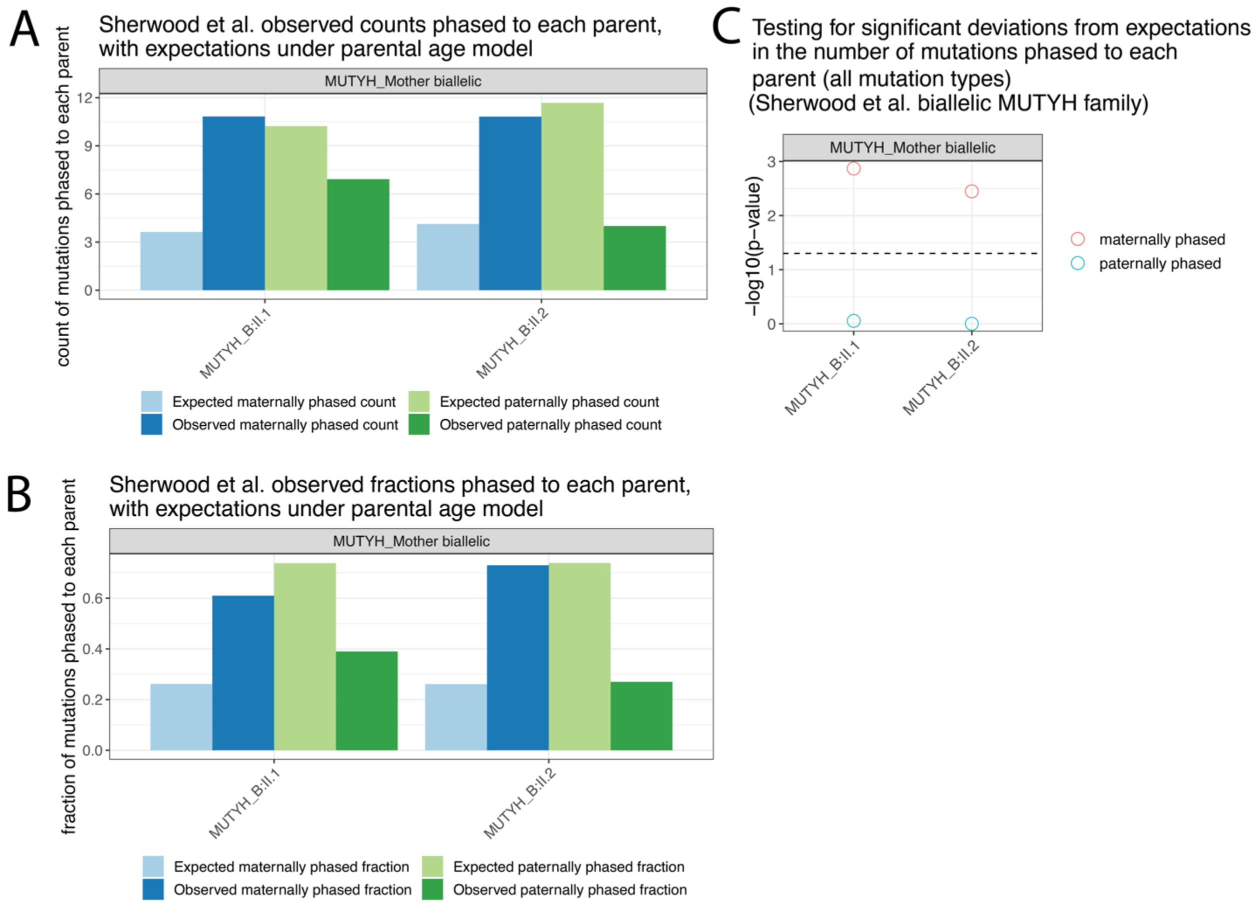


**Figure S13. Significantly more DNMs were phased to the biallelic *MUTYH* mother in Sherwood et al. (2023) in both of her children (*MUTYH*_B.II.1 and *MUTYH*_BII.2). A)** The counts of mutations phased to maternal (dark blue) and paternal (dark green) haplotypes reported by Sherwood et al. (2023), with expectations from the parental age model in light blue and light green. **B)** As in (A), but showing the fraction of phased mutations phased to each parent. Note the substantial elevations of maternally-phased mutations that they report. **C)** Under our significance testing threshold, both children of the biallelic *MUTYH* mother from Sherwood et al. show a significant elevation of overall mutations phased to the maternal haplotype than what is expected under the parental age model.

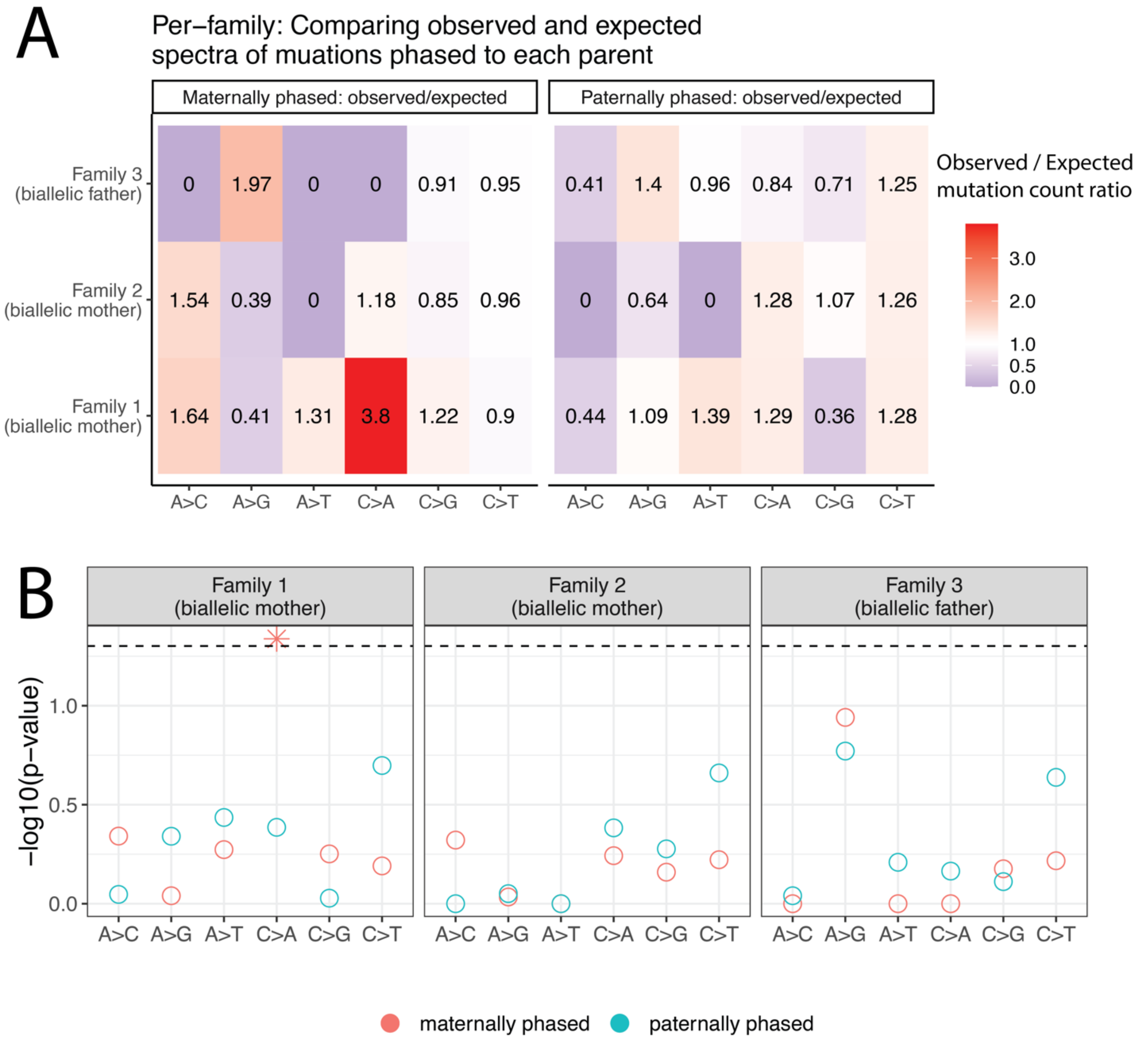


**Figure S14. Significantly more C>A mutations phased to the children of Family 1 (biallelic mother) than expected. A)** The ratio of observed/expected (under the parental age model (corrected for accessible genome size and individual phasing success rates) mutations phased to maternal and paternal haplotypes across the three biallelic *MUTYH* families in this study. **B)** The probability of observing greater than or equal to the number of mutations phased to each parent under the parental age model (corrected for accessible genome size and individual phasing success rates). Only Family 1 shows a significant result for C>A mutations phased to the carrier parent (biallelic mother).

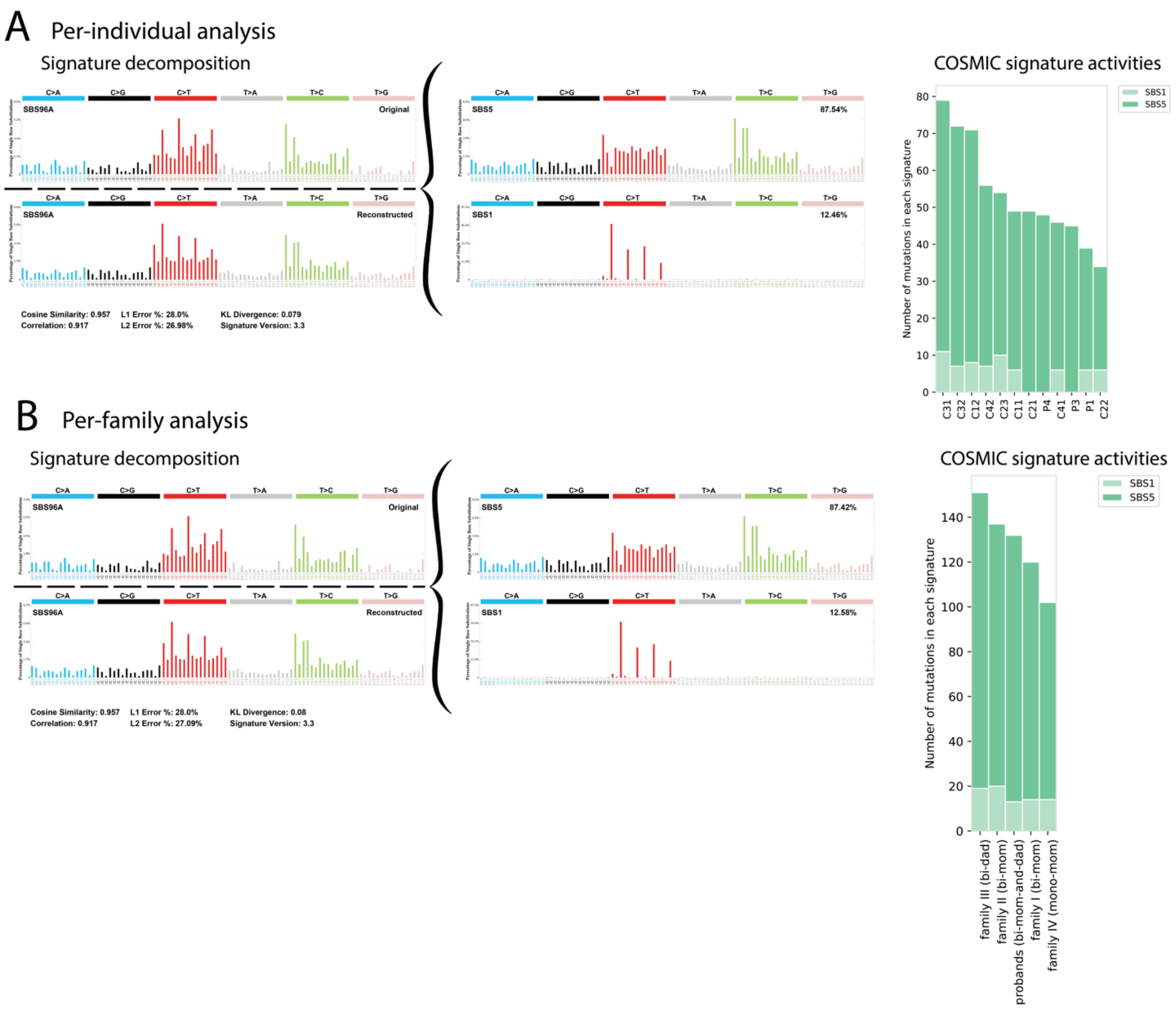


**Figure S15. Mutational signature extraction does not find activity of SBS18 or SBS36.** Results of SigProfilerExtractor, which extracts novel mutation signatures from the 3-mer mutation spectrum either at the per-individual level **(A)**, or across spectra summed across siblings within the same family **(B)**. The novel signature is then deconvoluted into known COSMIC signatures. The novel signature is shown in each row as "SBS96A (original)", and its reconstruction based on known COSMIC signatures is shown below ("SBS96A (reconstructed)"). The COSMIC signatures used to reconstruct the signatures are shown to the right of the brackets (SBS1 + SBS5). The cosine similarity reported is between the original and reconstructed signatures, indicating how well COSMIC signatures can be used to reconstruct the signatures extracted from the empirical data. The inferred activities (numbers of mutations contributed) of each signature is shown in the "COSMIC signature activities" plots. Analysis was repeated using COSMIC v2 in which signatures SBS18 and SBS36 are not separated, but did not yield qualitatively different results.

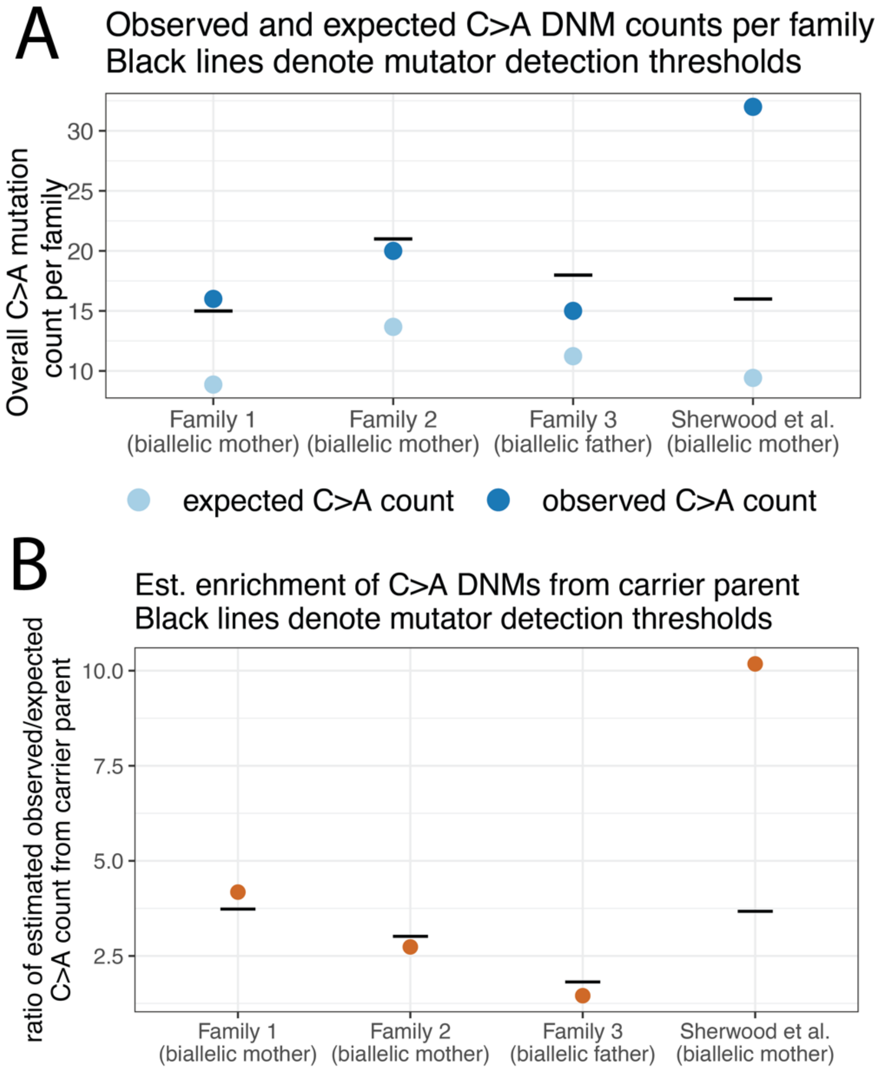


**Figure S16. Estimating the minimum *MUTYH* effect sizes that we have power to detect in the male and female germlines. A)** Observed (dark blue) and expected (light blue) C>A mutation counts in the children of each family with a biallelic parent. Horizontal black lines show the minimum number of mutations needed to reject the null parental age model ("mutator detection threshold"). Of the two biallelic mother families, Family 1 exceeds the threshold, implying a significant C>A mutation rate elevation while Family 2 falls just short, and Family 3 (biallelic father) falls further short. The family with a biallelic *MUTYH* mother from Sherwood et al. (2023) is included, and has a much more elevated C>A count than the families in this study. **B)** Estimates of the effect size of *MUTYH* on the number of C>A mutations transmitted by the carrier parent relative to expectations under the parental age model. Orange points indicate an estimate based on observed mutation counts in the children of each family, assuming all excess C>A mutations beyond the parental age expectations were inherited from the carrier parent. The horizontal black lines show the minimum effect size that exceeds a one-tailed 95% confidence interval above the Jónsson (2017) parental age model expectation (corresponding to the mutation counts denoted by the horizontal lines in (A)). These effect sizes represent estimates of the overall effect of *MUTYH* variants across gametes from the biallelic carrier parent. The minimal detectable effect size is much lower for Family 3 (biallelic father) than for Families 1 or 2 (biallelic mothers), as fathers transmit much higher numbers of mutations to their offspring, which makes it surprising that we detect significantly elevated C>A rates in Families 1 and 2 but not Family 3. This result suggests that *MUTYH* variation may exert a proportionally stronger effect on the female germline compared to the male germline. The large elevation of C>A mutations in the biallelic mother family from Sherwood et al. (2023) implies a higher effect size in the carrier parent than any seen in the families in this study. See **Figure S17** for this analysis based on per-individual mutation counts.

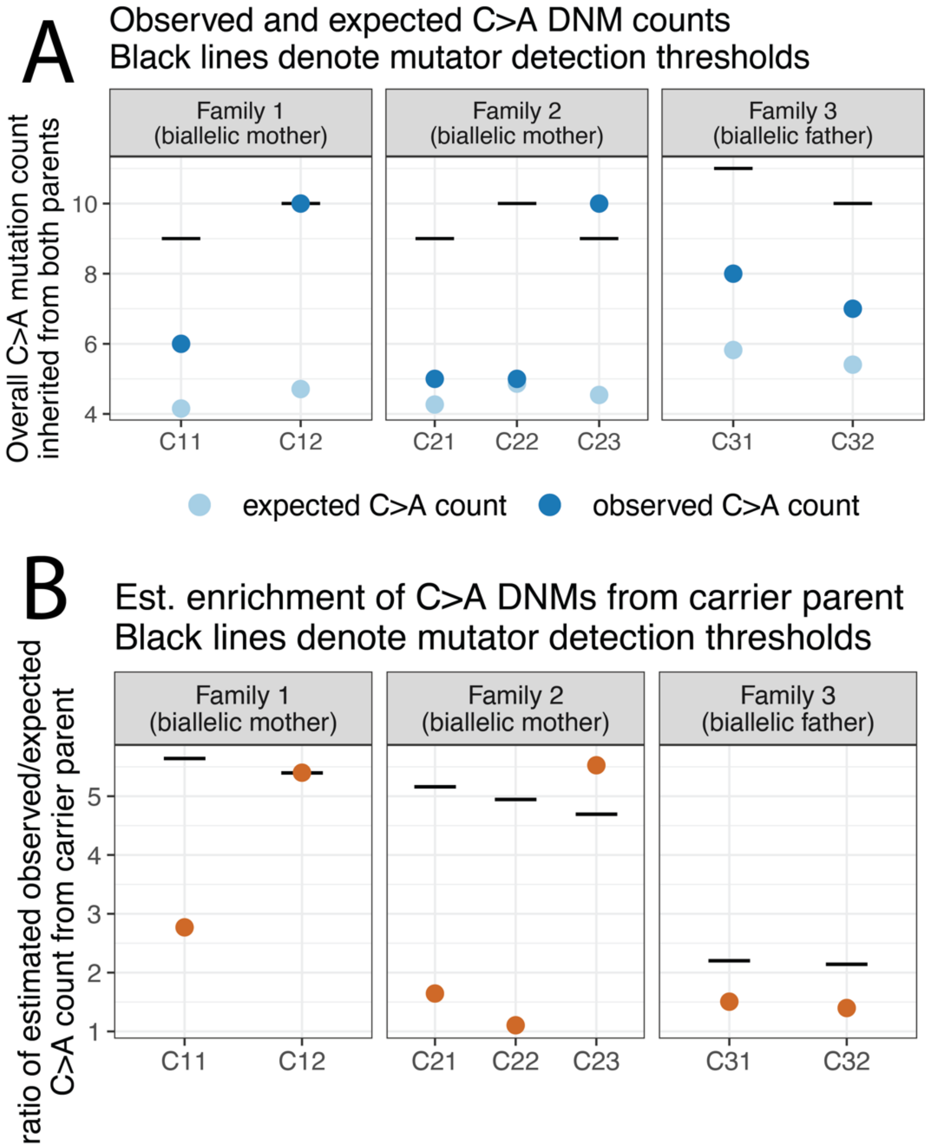


**Figure S17. Estimating the minimum per-individual *MUTYH* effect sizes that we have power to detect in the male and female germlines.** As in main text **Figure S16,** but based on individuals' C>A DNM counts rather than C>A DNM counts summed per family. **A)** Observed (dark blue) and expected (light blue) C>A mutation counts per individual in biallelic carrier parent families. Horizontal black lines show the number of mutations needed to reject the null parental age model ("mutator detection threshold"). As can be seen in **Figure 3C**, among the children of biallelic carriers, only C12 and C23 reach that threshold. **B)** Estimates of the effect size of *MUTYH* on the number of C>A mutations transmitted by the carrier parent relative to expectations under the parental age model. Orange points indicate an estimate based on observed mutation counts, if all excess C>A mutations beyond the parental age expectations are assigned to the carrier parent. The horizontal black lines show the minimum effect size that exceeds a one-tailed 95% confidence interval above the Jónsson (2017) parental age model expectation (corresponding to the mutation counts denoted by the horizontal lines in (A)). Note that the minimum detectable effect size in the children of the biallelic father is much lower than that of the children of biallelic mothers, as fathers transmit much higher numbers of mutations to their offspring. Despite this lower threshold, we observe no significant elevation of the C>A count in the biallelic father family.
